# Genome-wide association study of delay discounting in 134,935 individuals identifies novel loci and transdiagnostic associations across mental and physical health

**DOI:** 10.1101/2024.09.27.24314244

**Authors:** Hayley HA Thorpe, Renata B Cupertino, Shreya Reddy Pakala, Pierre Fontanillas, Mariela V Jennings, Jane Yang, John J Meredith, Tiffany Greenwood, Sevim B Bianchi, Laura Vilar-Ribó, Maria Niarchou, 23 andMe Research Team, Sarah L Elson, Trey Ideker, Lea K Davis, James MacKillop, Harriet deWit, Daniel E Gustavson, Travis T Mallard, Abraham A Palmer, Sandra Sanchez-Roige

## Abstract

Delay discounting (DD), a person’s preference for smaller immediate rewards over larger delayed rewards, is a heritable trait that is associated with psychiatric and physical outcomes, yet the biological mechanisms underlying these links are not known. We performed a GWAS of DD using 134,935 adults and identified 14 genome-wide significant loci. We observed genetic correlations between DD and 73 behavioral, physical, and neuroimaging traits, many of which persisted even after accounting for educational attainment, intelligence, and executive function. Network analysis revealed that the associations between DD and certain traits were explained by both overlapping and trait-specific biological processes. In a hospital-based cohort (*N* = 66,917), DD polygenic scores were associated with 212 medical conditions. These results demonstrate that DD has a pleiotropic and polygenic common variant architecture, and is genetically associated with numerous outcomes, making it a promising endophenotype for psychiatric and physical health.

## INTRODUCTION

Delay discounting (DD) is the tendency for individuals to devalue delayed rewards^1^. DD belongs to a broader family of traits that measure aspects of cognitive and executive function, including decision-making^2,3^ and impulsivity^4,5^. In humans, DD is typically measured using a hypothetical monetary temporal discounting task^1^ wherein participants make a series of choices between a smaller, immediate reward and a larger, delayed reward. These choices can be modeled as a hyperbolic discounting curve, which may be steeper (i.e., greater discounting of delayed rewards) or shallower. Steeper discounting is positively correlated with substance use disorders (**SUDs**)^6–8^, behavioral addictions (e.g., gambling disorder)^9^, body mass index (**BMI**)^10,11^, attention-deficit hyperactivity disorder (**ADHD**)^12^, bipolar disorder^10^, schizophrenia^10,13,14^, and post-traumatic stress disorder (**PTSD**)^15^. In contrast, shallower DD is observed in obsessive-compulsive disorder, certain personality disorders^16^, and anorexia nervosa^10,17,18^. Based on this spectrum of associations, DD has been proposed as a transdiagnostic marker of health, yet the mechanisms mediating these associations are poorly defined.

Several lines of evidence indicate that DD is heritable. Twin studies have estimated the heritability of DD to be anywhere from 35 and 62%^19,20^. More recently, our prior genome-wide association study (**GWAS;** *N*=23,217) estimated the SNP heritability of DD to be 12.2%, and showed positive genetic correlations between greater DD and ADHD, smoking initiation, BMI, as well as negative genetic correlations with schizophrenia^21^, educational attainment (*r_g_* = -0.93) and childhood IQ (*r_g_* = -0.63). The present study was designed to further characterize the relationship between DD and an array of psychiatric, physical, and cognitive measures. We built upon our prior DD GWAS^21^ by increasing the sample size by more than 5-fold to 134,935 US-based 23andMe, Inc. research participants of European ancestry. We performed global^22,23^ and local^24^ genetic correlations and conducted network analyses to identify specific biological processes shared between DD and various cognitive and health-related traits. Considering the strong genetic correlations between DD, educational attainment, and IQ^21^, we used multivariate techniques to parse the genetic contributions specific to DD from those shared with other cognitive traits. Finally, to further examine the link between genetic liability for DD and medical outcomes, we calculated DD polygenic scores (**PGS**) in a hospital-based cohort and conducted a polygenic phenome-wide association study (**PheWAS**).

## RESULTS

### GWAS of DD uncovered associations with 14 independent loci

We measured DD in US-based adult research participants (for participant demographics, see **Supplementary Table 1**) using the 27-item Monetary Choice Questionnaire (**MCQ**)^1^, which was administered via 23andMe’s user portal. DD was summarized using the temporal discounting value (***k***)^25^, wherein a larger value reflects a steeper discounting rate (i.e., a preference for immediate rewards over delayed gratification; see **Methods**, **Supplementary Table 2**). As in our prior GWAS, we log_10_ transformed *k* to better approximate a normal distribution. Only the participants who clustered within a European genetic ancestry panel (*N* = 134,935; hereafter referred to as “European cohort”) were used for GWAS (see **Supplementary Methods**). This cohort includes individuals from our prior publication^21^. We performed a GWAS using an additive genetic model that included age, sex, the first 5 genetic principal components, and indicator variables for genotype platforms as covariates (for single nucleotide polymorphism [**SNP**] quality control and inclusion, see **Supplementary Table 3**). Using Linkage Disequilibrium Score Regression (**LDSC**)^22,23^, we estimated the SNP-based heritability (***h***^2^ ) to be 9.85±0.57% (Intercept: 1.01±0.01). We identified 14 independent loci exceeding the genome-wide significance threshold of *P* < 5.00×10^−08^ (**Figure 1**; **Supplementary Tables 4-5**; see **Supplementary Figures 1-14** for LocusZoom plots).

**Figure 1.**
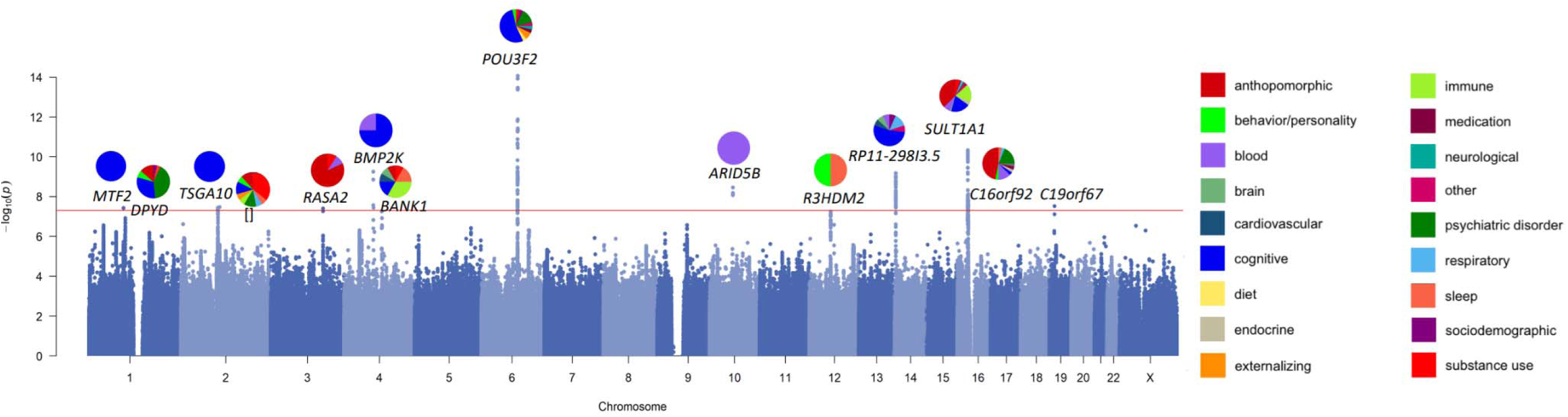
Manhattan plot shows 14 independent loci associated with DD. Pie plots show categories, highlighted using different colors, previously associated with loci (nearest gene annotated) from the GWAS catalog.

All lead SNPs except for rs57823886 (chr19p13.12) were located within loci that had prior published GWAS associations with at least 1 and up to 60 other traits, most of which were behavioral traits (**Figure 1**, **Supplementary Table 6**). For example, the strongest association (rs34645063, *P* = 3.20×10^−13^) was located on chr6q16.1 between the genes *MMS22L* and *POU3F2*. SNPs in linkage disequilibrium with rs34645063 have previously been implicated in a variety of GWAS including risk-taking behaviors^26,27^, substance use [i.e., alcohol consumption^28^, smoking initiation^28–31^, caffeine intake^32,33^], psychiatric disorders (e.g., bipolar disorder)^34–38^, externalizing psychopathology^39^, neuroticism/mood instability^40,41^, as well as educational attainment^42,43^, intelligence^44–48^, socioeconomic factors (household income^49^), and BMI^50–52^. Notably, we did not replicate our previously reported association with rs6528024 (chrXq13.3, *P* = 5.30×10^−02^)^21^.

### Functional annotation identified 93 DD candidate genes

To identify potential candidate genes associated with DD, we performed gene-(i.e., MAGMA^53^, H-MAGMA^54,55^) and transcriptome-based analyses (i.e., S-PrediXcan^56–58^, **Supplementary Tables 7-10**). MAGMA, which maps SNPs to genes based on physical proximity, identified 25 genes, 21 (84%) of these genes were located within 5 of the 14 GWAS loci (**Supplementary Table 7**). Next, we used H-MAGMA to incorporate GWAS results with chromatin interaction profiles from human brain tissues and iPSC lines, which implicated 66 unique genes across different cell-types (27.84% iPSC derived neurons, 24.74% midbrain dopamine neurons, 24.74% cortical neuron, 22.68% iPSC derived astrocyte) and developmental stages (50.82% fetal brain, 49.18% adult brain; **Supplementary Table 8**). Finally, we used S-PrediXcan to identify correlations between DD and predicted gene expression in the brain; this analysis identified 33 unique genes, 6 of which were consistently upregulated (i.e., *EIF3C*, *HAUS4*, *LYG1*, *NPIPB6*, *SULT1A2*) and 9 of which were consistently downregulated (i.e., *CCDC18*, *INO80E*, *NPIPB7*, *NPIPB9*, *RP11-1348G14.4*, *SH2B1*, *SULT1A1*) in more than one brain region (**Supplementary Table 9**). Overall, these analyses identified 93 unique genes associated with DD (**Supplementary Table 10**), of which 27 (29.03%) were identified by more than one method. Of these genes, 50 (53.76%) were located within or near the 14 loci identified by our GWAS; the most notable of these were the 18 (19.35%) genes within the ch16p11.2 GWAS locus.

### DD was globally genetically correlated with 73 psychiatric, cognitive, and physical health traits

We used LDSC^22,23^ to estimate genome-wide (or “global”) genetic correlations (***r_g_***) between DD and 109 other published traits, which were selected based on previously known phenotypic correlations (e.g., SUDs, impulsivity measures, metabolic traits). DD was genetically correlated with 73 of the 109 complex traits, including substance use, psychiatric disorders, impulsivity, cognition, physical health, brain measures, and sociodemographic variables (**Figure 2**, **Supplementary Table 11**).

**Figure 2.**
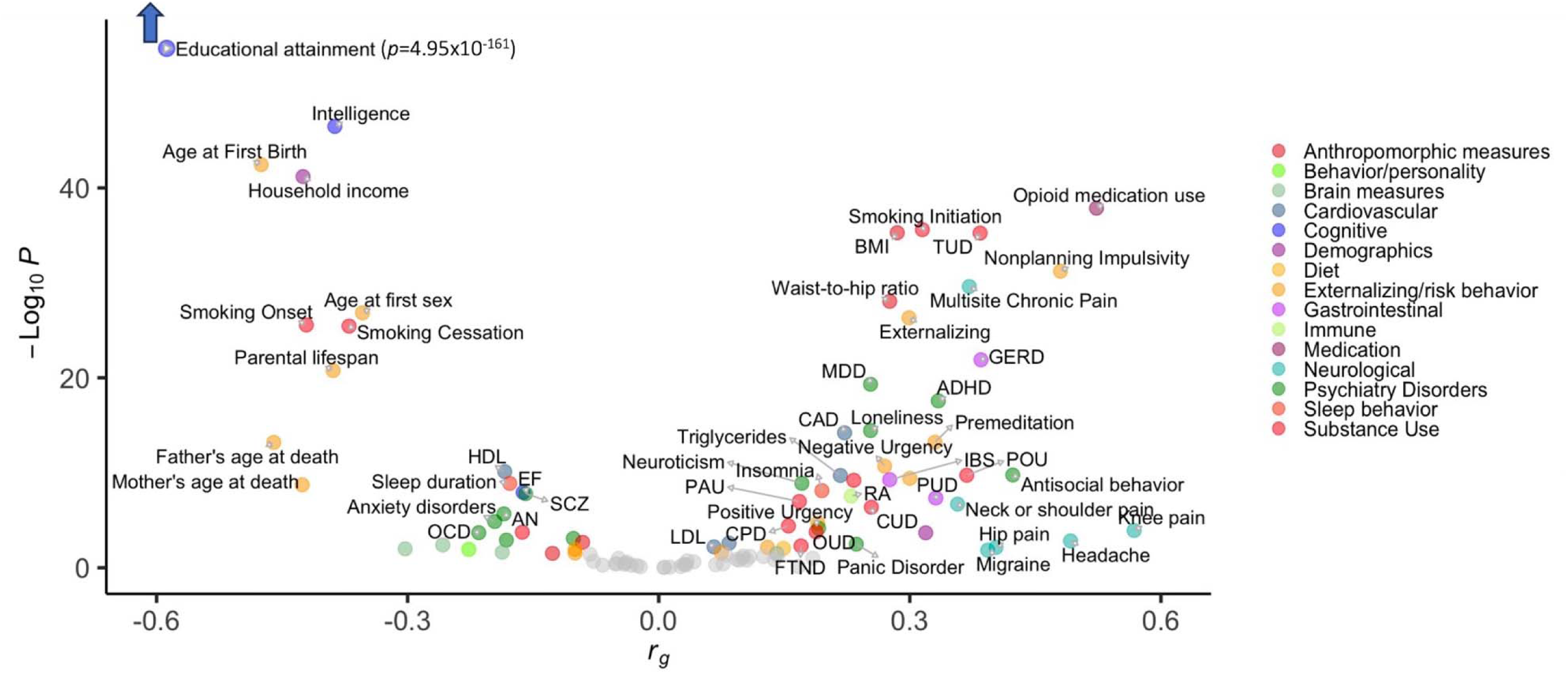
LDSC genetic correlations (*r_g_*) between DD and 109 traits from publicly available GWAS. 73 FDR-significant results are colored and labeled. See **Supplementary Table 11** for complete results.

#### Substance use and SUDs

DD showed positive genetic correlations with several substance use traits (e.g., smoking initiation *r_g_* = 0.32±0.02, problematic alcohol use *r_g_* = 0.37±0.06) and SUDs (e.g., tobacco use disorder [**TUD**] *r_g_* = 0.38±0.03, cannabis use disorder *r_g_* = 0.25±0.05, addiction risk factor *r_g_* = 0.23±0.04).

#### Psychiatric disorders

DD showed positive genetic correlations with some psychiatric conditions (i.e., antisocial behavior *r_g_*= 0.42±0.07, ADHD *r_g_* = 0.33±0.04, major depressive disorder *r_g_*= 0.25±0.04, loneliness *r_g_*= 0.25±0.03, panic disorder *r_g_* = 0.24±0.08, suicide attempt *r_g_* = 0.19±0.05), but negative genetic correlations with others (e.g., obsessive-compulsive disorder *r_g_* = -0.22±0.06, anorexia nervosa *r_g_* = -0.18±0.04, autism spectrum disorder *r_g_* = -0.18±0.06; schizophrenia *r_g_* = -0.16±0.03, bipolar disorder *r_g_* = -0.10±0.03).

#### Impulsivity and externalizing behavior

DD showed positive genetic correlations with some impulsivity facets (e.g., nonplanning impulsivity *r_g_* = 0.48±0.04, premeditation *r_g_* = 0.33±0.04) and small negative genetic correlations with others (i.e., sensation seeking *r_g_* = -0.10±0.04, perseverance *r_g_* = 0.10±0.05). We observed a positive genetic correlation with externalizing (*r_g_* = 0.30±0.03).

#### Cognition and executive function

DD showed negative genetic correlations with cognitive-related traits, including educational attainment (*r_g_* = -0.57±0.02), intelligence (*r_g_* = -0.39±0.03), and executive function (*r_g_* = -0.16±0.03). These traits included the strongest and most significant genetic correlations.

#### Physical health conditions

DD showed positive genetic correlations with pain-related traits (e.g., migraine *r_g_* = 0.39±0.16; multisite chronic pain *r_g_*= 0.37±0.03). We observed other positive genetic correlations between DD and digestive (e.g., gastro-esophageal reflux disease *r_g_* = 0.38±0.04, irritable bowel syndrome *r_g_* = 0.28±0.04), cardiovascular (e.g., coronary artery disease *r_g_*= 0.22±0.03), immune (i.e., rheumatoid arthritis *r_g_* = 0.23±0.04), sleep (e.g., insomnia *r_g_* = 0.19±0.03), and anthropometric variables (e.g., BMI *r_g_* = 0.29±0.02, waist-to-hip ratio *r_g_* = 0.28±0.02).

#### Brain measures

DD showed a positive genetic correlation with limbic network structural connectivity (*r_g_*= 0.14±0.07). There were negative genetic correlations with functional connectivity of limbic (*r_g_*= -0.30±0.12) and somatomotor (*r_g_* = -0.19±0.08) networks, and with intracranial volume (*r_g_*= -0.26±0.09).

#### Sociodemographics

DD was negatively genetically correlated with sociodemographic variables, such as household income (*r_g_* = -0.43±0.03) and parental lifespan (*r_g_*= -0.39±0.04).

### Local genetic correlation analysis revealed 3 pleiotropic hotspots, as well as 77 other loci of interest

To more finely pinpoint the specific loci contributing to global genetic correlations, we performed local analysis of [co]variant annotation (**LAVA**)^24^ across 2,495 semi-independent, approximately equally-sized (∼1Mb) genomic loci, which we downloaded from the LAVA website (https://github.com/cadeleeuw/lava-partitioning). Of the 73 globally genetically correlated traits, 36 traits showed local genetic correlations, which were dispersed across 80 loci (**Supplementary Figure 15**, **Supplementary Table 12**). The greatest number of overlapping loci were with intelligence (*N* = 12 loci).

We detected 3 pleiotropic loci or “hotspots” – chr6q16.1, chr3p21.31, and chr5q14.3 – where DD was locally genetically correlated with 5 or more traits (**Supplementary Figure 16**). Local versus global genetic correlations were not always consistent. At the chr6q16.1 locus (comprising 14 genes), which was the most pleiotropic hotspot, we observed consistent negative local correlations with educational attainment, intelligence, bipolar disorder, age at first birth, systolic blood pressure, infant head circumference at 6-30 months old, and executive function, all of which were consistent with the global genetic correlations. We also observed an inconsistent negative genetic correlation with externalizing, which showed a positive global genetic correlation. At the chr3p21.31 locus (127 genes), we identified positive genetic correlations with gastro-esophageal reflux disease, multisite chronic pain, externalizing, major depressive disorder, and ADHD, which 227 were consistent with the global genetic correlations, as well as circumference at birth, which 228 had shown a negative global genetic correlation. Finally, at the chr5q14.3 locus (10 genes), we observed 2 consistent positive genetic correlations with cannabis use disorder and loneliness, 2 consistent negative genetic correlations with executive function and intelligence, and 1 inconsistent positive genetic correlation with infant head circumference, which had shown a negative global genetic correlation.

Global genetic correlations may not reflect the strength of local genetic correlations if only a few loci are genetically correlated. For example, 9 traits showed local genetic correlations but no global genetic correlations with DD. 7 of these traits were locally genetically correlated with DD at only 1 or 2 loci (**Supplementary Figure 15**, **Supplementary Table 12**).

Finally, we found that DD was genetically correlated with 10 traits at only one locus per trait (**Supplementary Table 12**). For example, we found a single positive local genetic correlation between DD and addiction risk factor at the chr11q23.1 locus, which contains the *NCAM1-TTC12-ANKK1-DRD2* gene cluster.

### Network analysis identifies overlapping and distinct biological processes underlying the relationships between DD and other complex traits

To characterize the biological processes shared between DD and other traits, we propagated MAGMA-identified DD genes in HumanNet-XC^59,60^ (**Supplementary Figure 17**; **Supplementary Table 13**). HumanNet-XC is a gene interaction network that integrates gene co-expression, protein-protein interactions, and genetic interactions, to model human biology and disease. We selected 6 traits (BMI, schizophrenia, educational attainment, externalizing, addiction risk factor, and ADHD) as representative phenotypes from cluster analysis based on global genetic correlations (**Supplementary Figure 18**).

All traits except ADHD shared common biological processes with DD (**Figure 3**). DD shared the greatest number of biological processes with educational attainment (22), followed by BMI (17), externalizing (13), schizophrenia (6), and addiction risk factor (1). No single process was shared among all 5 traits. Rather, there was some specificity in the traits represented in different processes. Educational attainment and externalizing overlapped with DD on 11 processes. We found 3 metabolic processes shared between DD and BMI (i.e., “Cobalamin metabolic process”, “Glycosylceramide metabolic process”, “3’-phosphoadenosine 5’-phosphosulfate metabolic process”). These metabolic processes associated with DD were also shared with schizophrenia, externalizing, and educational attainment, highlighting the overlapping mechanisms that contribute to multiple traits simultaneously. Otherwise, there was some degree of specificity in DD biological processes shared only with one other trait, such as schizophrenia, externalizing, educational attainment, or BMI (but none exclusively shared with addiction risk factor; **Figure 3**).

**Figure 3.**
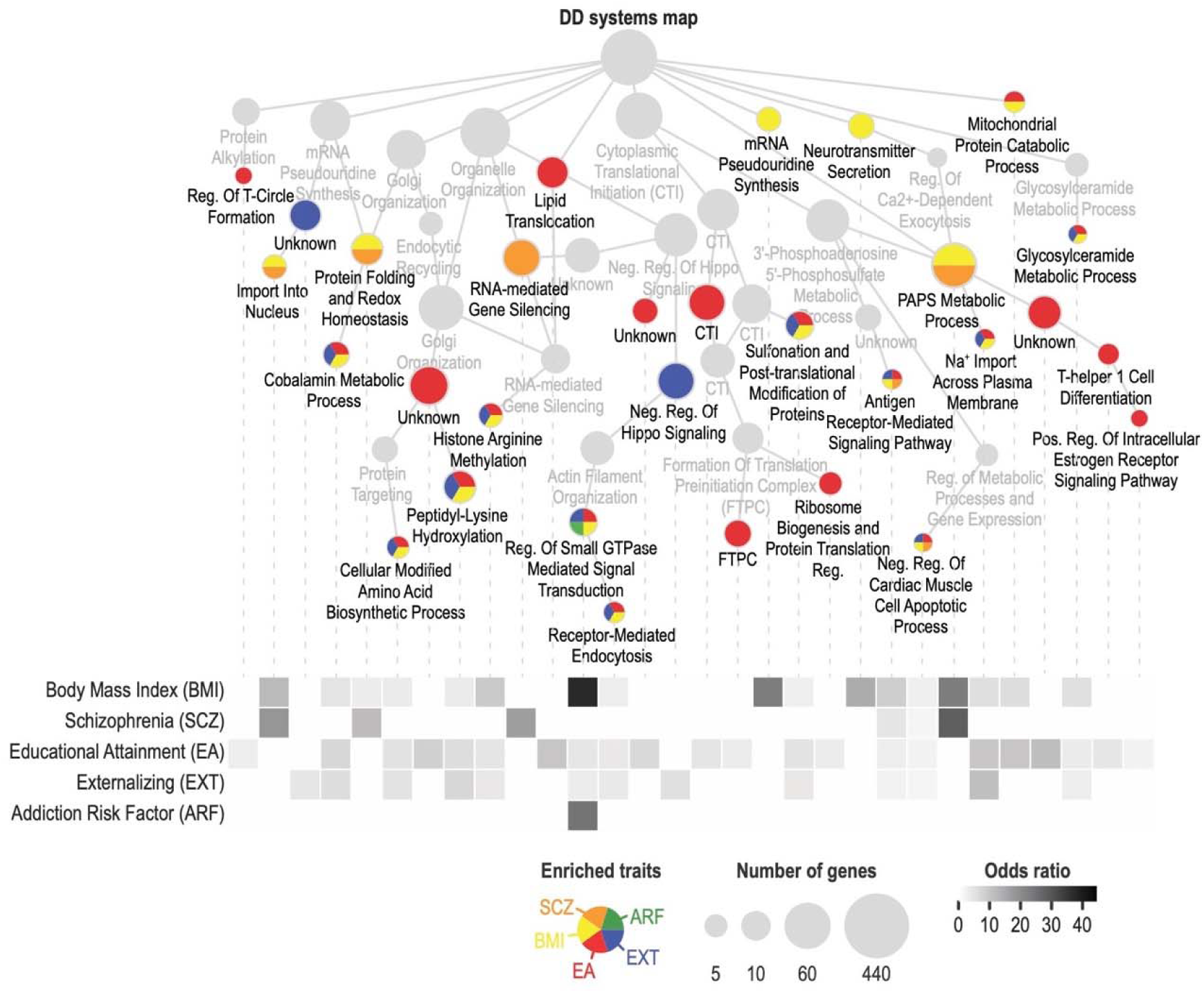
Biological processes shared across DD and related traits. Related traits include body mass index (**BMI**), schizophrenia (**SCZ**), educational attainment (**EA**), externalizing (**EXT**), addiction risk factor (**ARF**), and attention deficit hyperactivity disorder (**ADHD**; not shown). The DD systems hierarchy was generated by propagating MAGMA-identified genes in HumanNet-XC, which is a gene interaction network, clustered into gene communities at various resolutions, and annotated using Gene Ontology biological processes. The upper panel shows select systems enriched with at least one of the 6 traits listed above. The node size indicates the number of genes in each system, and the node color indicates the enriched trait(s). The lower panel shows a heatmap of odds ratio for DD and each of the 6 traits, calculated for each process shown in the upper panel. No processes implicated in DD were enriched with ADHD-associated genes. See **Supplementary Table 13** for full results.

### GWAS-by-subtraction identified persistent genetic correlations after accounting for educational attainment, intelligence, and executive function

DD is known to be negatively phenotypically correlated with cognitive traits like educational attainment and IQ^61–63^. Accordingly, our global and local genetic correlations both identified negative genetic correlations between cognitive traits and DD. We used the GWAS-by-subtraction^64^ approach within genomic structural equation modeling (**Genomic SEM**)^65^ to generate GWAS summary statistics (“*DDminusCognition*”) that excluded genetic variance related to educational attainment^42^ (*N* = 765,283), intelligence^46^ (*N =* 269,867), and executive function^66^ (*N* = 427,037; **Figure 4A**). While the *DDminusCognition* GWAS did not yield any significant loci, 6 out of the original 14 (42.86%) genome-wide significant loci remained significant at a suggestive threshold (*P* < 5.00×10^−05^; **Figure 4B**; **Supplementary Table 14**). GWAS signals were most attenuated by accounting for educational attainment, the GWAS with the largest sample size (Supplementary Figure 19).

**Figure 4.**
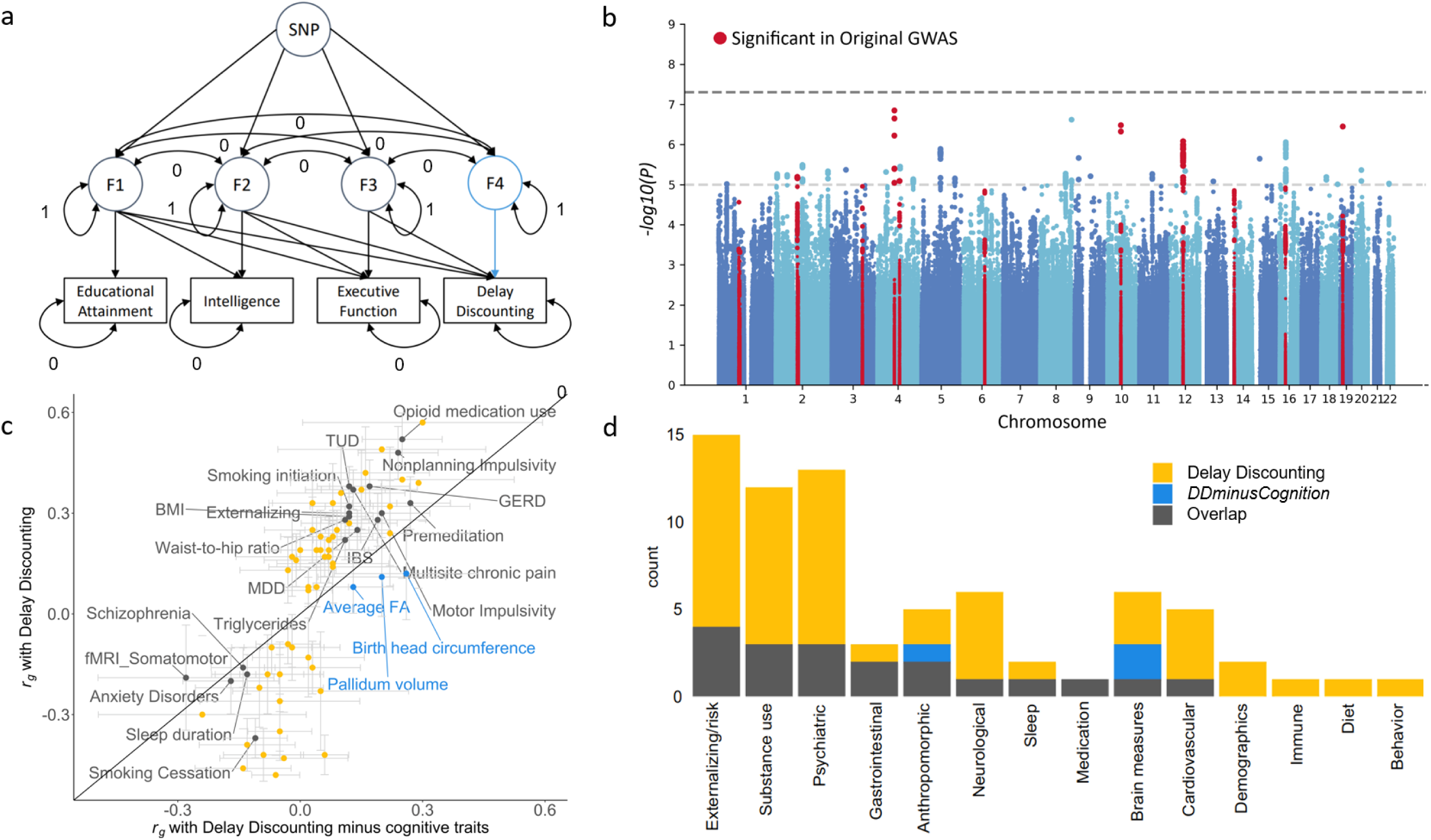
Parsing DD genetic variance from that of other cognitive traits revealed 19 of the 70 original genetic correlations persisted. **a)** Cholesky model structure showing path estimates for a single SNP fitted using genomic SEM. Educational attainment, intelligence, executive function, DD, and SNP are observed variables; F1-4 are latent (unobserved) variables. F4 represents *DDminusCognition*. The covariances between all the latent variables are constrained to 0. The residual variances of educational attainment, intelligence, executive function, and DD are constrained to 0, so that all variance is explained by the latent factors. The variances of the latent factors are constrained to 1. **b)** Manhattan plot for the *DDminusCognition* GWAS. The upper dashed line denotes the genome-wide significance threshold (*P* = 5.00×10^−08^) and lower dashed line indicates nominal significance threshold (*P* = 5.00×10^−05^). Red dots indicate SNPs that were significant in the original DD GWAS. **c)** DD genetic correlations (*r_g_*±standard error) exclusively significant in the original GWAS (yellow), the *DDminusCognition* GWAS (blue), or significant in both GWAS (gray; **Supplementary Table 16**). **d)** Number of significant genetic correlations per trait category that were exclusive to the original GWAS, the *DDminusCognition* GWAS, or significant in both.

We repeated the global genetic correlation analysis, and found that 19 (27.14%) of the 70 original genetic correlations with DD persisted after accounting for cognitive-related factors (**Figure 4C-D**; **Supplementary Table 15**); these include significant genetic correlations with a broad range of traits and categories, such as smoking (i.e., smoking initiation *r_g_* = 0.12±0.03, TUD *r_g_* = 0.12±0.04, smoking cessation *r_g_* = -0.11±0.04), impulsivity (i.e., premeditation *r_g_* = 0.27±0.05, nonplanning impulsivity *r_g_* = 0.24±0.05, motor impulsivity *r_g_* = 0.20±0.06), externalizing (*r_g_* = 0.12±0.04), psychiatric disorders (i.e., SCZ *r_g_* = -0.14±0.04, major depressive disorder *r_g_* = 0.14±0.04), anthropomorphic measures (i.e., BMI *r_g_*= 0.29±0.02, waist-to-hip ratio *r_g_*= 0.28±0.02), brain measures (i.e., somatomotor functional connectivity (*r_g_*= -0.19±0.08), gastrointestinal traits (i.e., gastro-esophageal reflux disease *r_g_* = 0.17±0.05, irritable bowel syndrome *r_g_* = 0.19±0.06), triglycerides (*r_g_* = 0.22±0.03), prescription opioid use (*r_g_* = 0.52±0.04), multisite chronic pain (*r_g_*= 0.37±0.03), and sleep duration (*r_g_*= -0.18±0.03). However, many other substance use (e.g., age of smoking initiation, opioid use disorder), psychiatric (e.g., ADHD, bipolar disorder, anorexia nervosa, obsessive-compulsive disorder), externalizing-related traits (e.g., age at first birth, age at first sex), neurological (e.g., migraine, hip pain), and cardiovascular (e.g., coronary artery disease) traits, among others, were not genetically correlated with DD following adjustment for cognitive-related factors (**Figure 4C-D**). Educational attainment had the largest impact on the change in genetic correlation between the original and GWAS-by-subtraction DD GWASs (**Supplementary Table 15**).

### DD polygenic score was associated with 212 medical outcomes

To extend the associations with DD to medical outcomes, we performed a PheWAS of DD polygenic scores (**PGS**) against 1,318 diagnostic outcomes in the BioVU medical cohort (*N_European_* = 66,917; *N_African_*= 12,383). In the European cohort, DD PGS was associated with 212 medical outcomes. We identified expected associations with 2 substance-related diagnoses (i.e., TUD OR = 1.14, CI 95%: 1.11-1.16; substance addiction and disorders OR = 1.09, CI 95%: 1.05-1.13) and 4 psychiatric disorders (e.g., mood disorders OR = 1.04, CI 95%: 1.02-1.06). We also found associations with 24 respiratory (e.g., chronic airway obstruction OR = 1.13, CI 95%: 1.10-1.16), 17 sense organs (e.g., myopia OR = 0.90, CI 95%: 0.85-0.95), and 36 circulatory (e.g., ischemic heart disease OR = 1.04, CI 95%: 1.01-1.07) conditions. Consistent with our genetic correlation and network analyses, we also found that DD PGS associated with 18 endocrine/metabolic conditions (e.g., Type 2 diabetes OR = 1.10, CI 95%: 1.08-1.12, obesity OR = 1.07, CI 95%: 1.05-1.10; **Figure 5**, **Supplementary Table 16**). Considering that we identified robust associations with smoking, we performed a sensitivity analysis controlling for TUD diagnosis. Some of the associations were largely persistent (e.g., sense organs, dermatologic traits, neoplasms) whereas other associations were attenuated (e.g., circulatory, respiratory, endocrine/metabolic) or nullified (e.g., psychiatric disorders; **Supplementary Figure 20**, **Supplementary Table 16**).

**Figure 5.**
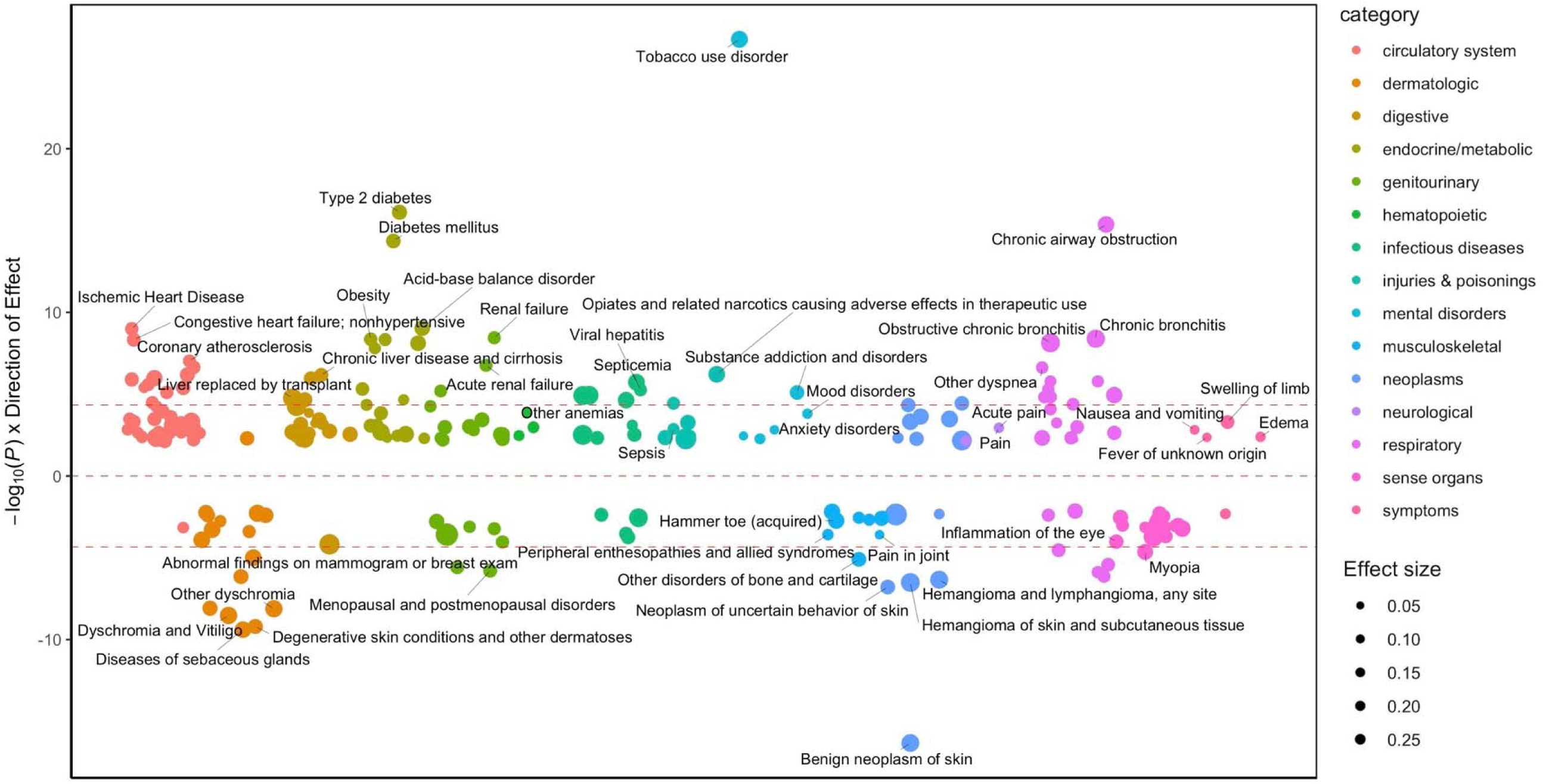
DD PGS PheWAS across 1,318 medical outcomes (*N_European_* = 66,917) in BioVU. Only FDR-significant results with 212 medical outcomes are annotated (see **Supplementary Table 16** for full results). Red dotted lines show the Bonferroni significance threshold.

There was some specificity in DD PGS associations across age groups (**Supplementary Table 16**). In the 19-25 age group (*N* = 6,241-7,246), we identified 2 associations, including positive associations with pregnancy complications (i.e., early or threatened labor; hemorrhage in early pregnancy OR = 1.30, CI 95%: 1.19-1.41, other conditions or status of the mother complicating pregnancy, childbirth, or the puerperium OR = 1.30, CI 95%: 1.18-1.42). In the 41-60 age group (*N* = 26,269-31,381), we identified 133 associations, including positive associations with substance use and psychiatric disorders (e.g., TUD OR = 1.17, CI 95%: 1.14-1.21, MDD OR = 1.09, CI 95%: 1.04-1.14) and physical health conditions (e.g., diabetes mellitus OR = 1.12, CI 95%: 1.09-1.15, obesity OR = 1.11, CI 95%: 1.07-1.15, renal failure OR = 1.11, CI 95%: 1.07-1.14). In the 61-100 age group (*N* = 22,405-26,855), we identified 44 associations, primarily positive associations with cardiovascular conditions (25.00% of all associations; e.g., myocardial infarction OR = 1.11, CI 95%: 1.06-1.15, ischemic heart disease OR = 1.06, CI 95%: 1.03-1.09); the only psychiatric association in this age group was with TUD (OR = 1.12, CI 95%: 1.08-1.17). We did not observe age-specific associations in the 0-11 (*N* = 7,042-8,459), 12-18 (*N* = 5,886-6,918), and 26-40 groups (*N* = 13,261-15,455) perhaps due to lower power due to smaller sample sizes.

We did not find any significant PheWAS associations in the African cohort (**Supplementary Table 17**), presumably due to both lower statistical power (*N*= 7,081-12,308) and the poor performance of our PGS, which was based on a European cohort. Portability of PGS decreases with greater genetic distance from the discovery sample, which is a well-known limitation^67^.

## DISCUSSION

We expanded our prior GWAS by over 5-fold^21^, leading to the identification of 14 independent loci and 93 candidate risk genes associated with DD. DD was genetically correlated with numerous psychiatric, neurobiological, and physical health outcomes; we identified 3 pleiotropic hotspots and biological processes underlying these associations. Although DD is an aspect of cognition, multivariate analyses identified genetic variation in DD that is not explained by broader cognitive-related measures. In a hospital cohort, DD PGS was associated with a multitude of medical outcomes, many of which may be related to health consequences of disadvantageous behaviors related to DD. This work pinpoints neurobiological targets of DD and sets the foundation for future studies that may enable the discovery of better prevention, diagnosis, and treatment mechanisms for a host of conditions.

The increased power from our current GWAS^21^ allowed us to identify 14 novel loci and 93 genes associated with DD. We identified 4 genes that were implicated by all 3 gene- and transcriptome-based methods: *SULT1A1*, *SH2B1*, *TUFM*, and *NPIPB6*. These genes are highly pleiotropic; they have been implicated in risk-taking^26^, substance use^28,68,69^, intelligence and cognitive ability^45,70^, obesity and BMI^52,68,71–82^, and brain morphometrics^83–85^. *SULT1A1* encodes the SULT1A1 sulfotransferase enzyme that is induced by dopamine in neuroepithelial cells *in vitro*^86,87^; the role of dopaminergic neurotransmission on DD is well-studied^88,89^. *SH2B1* encodes the SH2B1 adaptor protein, which mediates the activation of various kinases and is implicated in neurite outgrowth^90–93^, neuron differentiation and brain growth^91,94^, and obesity-related phenotypes^95–98^. Recent GWAS^94^ and candidate gene^94,99^ studies implicated *SH2B1* variants in fluid intelligence, aggression, and brain growth, which are partially corroborated by deletion of *Sh2b1* in mice^94,99^. *TUFM* encodes the Tu Translation Elongation Factor, Mitochondrial protein; it is hypothesized that proteins maintaining mitochondrial synthesis like TUFM mediate synapse development, function, and plasticity that ultimately impact cognition and executive functions^100^. The function of *NPIPB6* is not defined, though one transcriptome-wide association study found that its downregulation in the basal ganglia is associated with intelligence-associated variants^101^. These four genes are concentrated in the chr16p11.2 locus, which has been implicated in many of the traits associated with DD, including psychiatric disorders (e.g., autism spectrum disorder, ADHD, schizophrenia, bipolar disorder)^102^, BMI and eating behaviors^103–107^, head size and brain volume^105–108^, and intellectual and cognitive ability^104,106,109^, most notably inhibitory control^105^.

We found that DD is genetically correlated with a constellation of other traits from multiple categories. This included genetic correlations with brain-related traits that are consistent with neuroimaging studies showing associations between limbic and somatomotor connectivity and DD in adults^110–113^ and children^114^. These host of genetic correlations may arise from unique combinations of gene sets and processes that are trait-specific, as reflected in our network analysis. We find similar broad-ranging associations between DD PGS and psychiatric (e.g., addiction, mood) and physical (e.g., cardiovascular, endocrine/metabolic) outcomes, many of which could be the result of behaviors related to DD, such as smoking. These findings mirror associations found across decades of epidemiological studies^8,14,63,115,116^. One of our most consistent observations was that DD was genetically correlated with various smoking traits, from aspects of smoking initiation to cessation. Prior studies indicate that steeper DD associates with relapse, especially for smoking^117^, which could suggest that pharmacologically or behaviorally targeting DD could enhance SUDs treatment response^118–120^, although we were unable to identify pharmacotherapies that target genes implicated in DD via drug repositioning (**Supplementary Tables 18-19**). We also noted that liability for steeper DD was not always disadvantageous. For example, we identified *negative* genetic correlations with obsessive-compulsive disorder, anorexia nervosa, autism spectrum disorder, schizophrenia, and bipolar disorder. Likewise, we identified negative associations between DD PGS and some dermatological, musculoskeletal, and sense organ conditions.

To date, the etiological factors underlying the association of DD with psychiatric and physical health outcomes are not known. Here, we report local genetic correlations provide candidate regions contributing to pleiotropy. We identified a hotspot on chr6q16.1, in which we observed positive and negative genetic correlations between DD and 8 psychiatric, cognitive, and physical traits. In other instances, we observed traits that were globally genetically correlated with DD but showed only one local genetic correlation. For example, the positive global genetic correlation with addiction risk factor was recapitulated at the chr11q23.1-2 locus that includes the *NCAM1-TTC12-ANKK1-DRD2* gene cluster, which is well-known for its association with reward processes and psychiatric conditions, including substance use disorders^121–132^. DD hotspots often span broad regions encompassing dozens of genes; DD can be measured in non-human animals^133–135^, which can be used to validate and parse some of these complex findings.

DD is part of a family of cognitive^61–63^ and executive function processes, including impulsivity^5^. While we observed genetic correlations between DD and educational attainment, intelligence and executive function, as well as several impulsivity facets, these constructs are dissociable at a genetic level. For example, cluster analysis identified that DD loaded onto a factor discrete from other impulsivity measures^136–139^, and we found through genetic correlation, network, and GWAS-by-subtraction analyses that DD only partially overlaps with educational attainment, intelligence, and executive function. Educational attainment was most strongly genetically correlated with DD. Although this may be explained in part by increased statistical power with the larger sample size, it is also consistent with the idea that educational attainment greatly requires shallow DD (e.g., working toward delayed rewards and long-term goal-setting^61^) or other non-cognitive factors that are negatively genetically correlated with DD^64^.

DD PGS PheWAS corroborated numerous positive associations with health outcomes observed by prior phenotypic studies^6,9,10,12,13,16–18^ that were also consistent with our genetic correlations. Age-stratified PheWAS revealed associations that were missed in the original analysis using the full cohort (e.g., pregnancy complications in the 19-25 group), but overall effect sizes for PheWAS associations were largely consistent across age groups, in line with prior observations that DD expression and heritability stabilizes by adolescence^2,19,20,140^. The persistence of some DD PGS associations with these outcomes, even after accounting for the co-occurrence of other adverse health conditions related DD (e.g., adjusting for TUD), asserts that many of these relationships are influenced by both biological and behavioral factors.

Our findings have several limitations. First, there may be a cohort bias in discounting rates since 23andMe research participants are a self-selected group that must first join 23andMe and then agree to participate in an uncompensated research study. These research participants tend to be more educated, older, and have a higher socioeconomic status compared to the general population^141^. We included age as a covariate in our analyses but not educational attainment^42^, since including heritable traits as covariates may introduce collider bias^142^. Education was accounted for in the GWAS-by-subtraction analysis, albeit at a substantial loss in power to detect significant loci unique to DD. Second, these analyses do not address causality between DD and other traits, which may be complex and bidirectional in some cases. For example, substance use has been shown to increase DD, yet steeper discounting rates also predict substance use^6^. We suspect SUDs play a minimal role on DD in our population because the 23andMe research cohort that we studied did not heavily engage in substance use^138,143^. Our investigations are correlational in nature and may be influenced by many other unmeasured factors, such as cross-trait assortative mating^144^; future studies using causality inference techniques and longitudinal analyses could clarify the direction of these associations. Finally, GWAS were only conducted in individuals with European genetic similarity due to the large sample size required to detect the small effect of individual variants, and future analyses should diversify genetic analyses as larger non-European samples become available.

DD is a fundamental cognitive process involved in daily life that is broadly related to psychopathology and can be easily evaluated in health and clinical populations^2^. Our findings show that DD shares overlapping biological underpinnings with numerous psychiatric, cognitive, and physical outcomes, which may help propel prevention and treatment strategies across a broad spectrum of human health.

## METHODS

### Sample and DD Assessment

All individuals included in this study were research participants from 23andMe, as previously described^21,139^. Participants provided informed consent and participated in the research online, under a protocol approved by the external Association for the Accreditation of Human Research Protection Programs (**AAHRPP**)-accredited Institutional Review Board (**IRB**), Ethical & Independent (**E&I**) Review Services (http://www.eandireview.com/). As of 2022, E&I Review Services is part of Salus IRB (https://www.versiticlinicaltrials.org/salusirb).

Participants completed the online 27-item MCQ as part of a larger survey to assess preference for smaller, immediate rewards versus larger, delayed rewards (see **Supplementary Methods** for questionnaire). The overall response pattern was used to derive temporal discounting functions (***k***), wherein higher values indicate a greater devaluation of delayed rewards (i.e., preference for immediate gratification). Participants with low response concordance or inappropriate response rates were excluded from the analysis (**Supplementary Methods**). *k* was not normally distributed; all further analysis was conducted on log10(*k*) values^25^.

### Genome-wide association study

We borrowed population descriptors recommended in a recent report by the National Academies of Sciences, Engineering, and Medicine to define our study cohorts^145^. GWAS included 134,935 unrelated 23andMe US-based participants with at least 97% European ancestry, as determined through an analysis of local ancestry (**Supplementary Methods**). DNA extraction and genotyping were performed on saliva samples at clinical laboratories of Laboratory Corporation of America, certified by the Clinical Laboratory Improvement Amendments (**CLIA**) and accredited by the College of American Pathologists (**CAP**). Quality control, imputation, and genome-wide analysis were conducted by 23andMe (**Supplementary Methods**), as previously described^21^. A total of 14,137,232 SNPs passed the GWAS quality control (**Supplementary Table 3**). Association tests were conducted via linear regression under an additive model using a proprietary pipeline developed internally by 23andMe (*P* < 5.00×10^−8^). We included age (inverse-normal transformed), sex, the top five principal components of genotype, and indicator variables for genotype platforms as covariates. We identified previous associations with the SNPs linked to DD based on GWAS catalog information^146^.

### Gene- and transcriptome-based analyses

We conducted bioannotation and bioinformatic analyses to further characterize the loci identified by the DD GWAS. Gene-set analysis was conducted using Multi-marker Analysis of GenoMic Annotation (**MAGMA**; v1.08)^53^ through the FUMA (v1.3.6a) web-based platform^147^, with significance determined at a Bonferroni threshold based on the total number of genes tested (*P* < 2.53×10^−6^). We used Hi-C coupled MAGMA (**H-MAGMA**)^54,55^ to assign intergenic and intronic SNPs to genes based on their chromatin interactions based on 4 Hi-C datasets. We applied a Bonferroni correction based on the total number of gene-tissue pairs tested (*P* < 9.78×10^−7^). We used S-PrediXcan to identify expression quantitative trait loci-linked genes across 13 brain tissues associated with DD. We applied a Bonferroni threshold based on the number of genes per tissue (*N* = 2,532-6,744; *P* < 1.97×10^−5^ to 7.41×10^−6^). See **Supplementary Methods** for more details on gene- and transcriptome-based analysis.

### Heritability and genetic correlation analyses

We used LDSC^22,23^ to calculate SNP-based heritability from common SNPs mapped to HapMap3 data^148^ using pre-computed linkage disequilibrium scores (“eur_w_ld_chr/”). We also used LDSC to estimate genetic correlations between DD and 109 other traits. Traits were selected based on previously known phenotypic associations between DD and related traits (e.g., SUDs, impulsivity measures, metabolic traits). LDSC estimates the genetic correlation between complex traits or diseases by leveraging summary statistics from GWAS and patterns of linkage disequilibrium across the genome. We applied a 5% FDR threshold to account for multiple testing.

### Local genetic correlation

LAVA^24^ was used to estimate the “local” *r_g_* between DD and the 109 traits used in LDSC. LAVA splits the genome into 2,495 non-overlapping blocks of approximately equal size (∼1Mb) while minimizing the LD between them and estimates the bivariate SNP genetic correlation between 2 phenotypes for each block. LAVA first runs univariate tests to determine the amount of local genetic signal for all traits of interest and filters out loci with non-significant heritability for each trait. It then performs bivariate tests to obtain the local genetic correlation between pairs of traits with significant heritability (*P* < 2.00×10^−5^). We further corrected the bivariate analysis using FDR correction.

### Network propagation and visualization

We modified the Python package NetColoc^149–151^ for network propagation of a single gene set. The genes associated with DD in MAGMA analysis were used as ‘‘seed’’ genes. For network propagation, we used a Random Walk with Restart algorithm^152^, simulating heat spreading from the “hot” seed genes to adjacent genes in the network. The total heat within the system is conserved by dissipating a constant fraction of heat from each gene with each iteration. The process of heat diffusion stabilizes at a steady-state solution, as detailed in Equation 1:

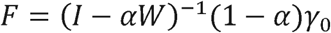

Where F represents the final heat distribution across nodes, Y_0_ is the vector of seed genes, W is the normalized adjacency matrix (HumanNet), and α (between 0 and 1) is the heat dissipation rate. After network propagation with α = 0.5, a network proximity score (**NPS**) was calculated for each gene by comparing the observed results to the expected results. The expected results were generated by propagating 100 random seed gene-sets, each of which was sampled to preserve the size and degree distribution of the original seed gene-set^153^. For each gene g, NPS was calculated as a Z score, comparing its observed heat F_g_,S after propagation to the average expected heat F_g_, and from the random sets, as outlined in Equation 2:

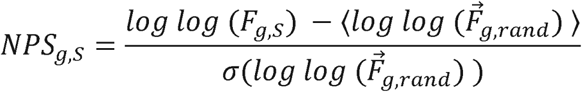

Where □□ denotes the mean of a vector, and denotes the standard deviation of a vector. All heat values are log-transformed to ensure they follow a normal distribution. To select for genes highly proximal to seed genes, we filtered for those with NPS > 3.

We built a multiscale DD network map using the Hierarchical community Decoding Framework (HiDeF) algorithm from Cytoscape’s CDAPS package^154^. HiDef employs persistent homology for simultaneous multi-scale community detection. The interaction network is translated into a fully connected similarity network, and then network communities are detected at various modularity levels, from smaller to larger communities by varying resolutions. Communities consistently appearing across resolutions were determined using Jaccard similarities and subsequently organized hierarchically, with their relationships assessed by the containment index (**CI**), which quantifies the overlap between communities v and w: If CI (v, w) exceeds the threshold, an edge is added from v to w, and any redundant edges are reduced in the final hierarchy. For the DD systems map, we used HiDeF with the maximum resolution at 5, with all other parameters set at their default values. The resulting communities were first annotated based on Gene Ontology (**GO**) Biological Processes using Enrichr^155^ and then manually refined. Gene enrichment for educational attainment, addiction risk factor, schizophrenia, externalizing, BMI, and ADHD was evaluated for each community using a hypergeometric test, followed by FDR correction. All hierarchical and network diagrams were created with *Cytoscape*^156^.

### GWAS-by-subtraction

We used the genomic SEM (ver0.0.5) R package^65^ and published GWAS of educational attainment^42^ (*N*= 765,283), intelligence^46^ (*N*= 269,867), and executive function^66^ (*N*= 427,037) to estimate SNP associations with DD that are independent of associations with cognitive-related traits. The model regressed GWAS summary statistics of educational attainment, intelligence, executive function, and DD onto 4 latent factors (F1-4), which were restrained to be uncorrelated (**Figure 4a**). Latent factors were regressed on each SNP and a GWAS of F4, which represents the genetic effects on DD that remained after accounting for the other traits (“*DDminusCognition*”), was conducted. We computed the genetic correlations within genomic SEM wherein a similar model as mentioned above was fitted, except the latent factors were regressed on the target trait instead of each SNP. This yielded a genetic correlation between the target trait and the variance in DD that is not overlapping with other cognitive-related traits.

### Phenome-wide association study

We conducted a PheWAS with DD PGS in the unrelated participants of the BioVU cohort (N_European_ = 66,917; N_African_ = 12,383) who provided electronic health records (**EHR**) to the Vanderbilt University Medical Center (**VUMC**; IRB #160302, #172020, #190418)^157^. We computed PGS for DD using PRS-Continuous shrinkage software (PRS-CS)^158^ using default settings to estimate shrinkage parameters and random seed fixed for reproducibility. For 1,817 medical conditions available through BioVU, we defined cases as patients who received at least two International Disease Classification (**ICD**, ICD-9 or ICD-10) diagnostic codes (also known as “phecodes”) in their EHR, and controls as patients with no diagnostic codes for that condition. To map ICD codes to phecodes, we used Phecode Map 1.2. Logistic models were fitted for each of the phecodes using the PheWAS v0.12 R package^159^ adjusting for sex, median age of longitudinal EHR, and the first ten principal components. A minimum of 100 cases were required for phecode inclusion, leaving a total of 1,318 phecodes. Sensitivity analyses were conducted using TUD status (phecode 318) as a covariate. To examine potential age-specific associations with health outcomes, independent PheWASs were conducted across 6 distinct age bins: 0-11, 12-18, 19-25, 26-40, 41-60, and 61-100. We used a 5% FDR threshold to correct for multiple testing.

### Drug Repositioning

We used DRUGSETS^160^ to facilitate genetically informed drug repositioning through drug gene-set analysis using MAGMA^53^. The data for drug-gene targets and interactions were drawn from the Clue Repurposing Hub and the Drug Gene Interaction Database. Gene-sets were created for 1,201 drugs comprising genes whose protein products are targeted by or interact with that specific drug. Competitive gene-set analysis was performed using the MAGMA software tool while conditioning on a gene-set of all drug target genes in the data (*N* = 2,281) to assess whether significant associations were due to effects unique to drug pathways or common properties of drug target genes. A Bonferroni significance threshold was applied based on the number of drug-gene sets analyzed (*N* = 735; *P* < 6.80×10^−5^). Additionally, drug gene-sets were categorized based on Anatomical Therapeutic Classification (**ATC**) III code, clinical indication, and mechanism of action. Each drug gene-set group with 5 or more drugs within the ATC (*N* = 63), mechanism of action (*N* = 51), and clinical indication (*N* = 80) categories was tested for enrichment of DD associated genes using a Bonferroni significance threshold (ATC III *P* < 7.94×10^−4^; clinical indication *P* < 6.25×10^−4^; mechanism of action *P* < 9.80×10^−4^).

## DATA AVAILABILITY

We will provide 23andMe summary statistics for the top 10,000 SNPs upon publication. The full 23andMe GWAS summary statistics will be made available through 23andMe to qualified researchers under an agreement with 23andMe that protects the privacy of the 23andMe participants. Please visit https://research.23andme.com/collaborate/#dataset-access/ for more information and to apply to access the data.

The following resources were used for secondary analysis: FUMA (https://fuma.ctglab.nl/), Ensembl build 85 (https://www.ebi.ac.uk/about/news/updates-from-data-resources/ensembl-version-85/), 1000 Genomes Project phase 3 (https://internationalgenome.org/data-portal/sample), Msigdb v.7.0 (https://data.broadinstitute.org/gsea-msigdb/msigdb/release/7.0/), Genotype–Tissue Expression (GTEx) v.8 project database (https://www.gtexportal.org/), H-MAGMA software and Hi-C datasets (https://github.com/thewonlab/H-MAGMA), PredictDB Data Repository (http://predictdb.hakyimlab.org/ and http://predictdb.org/), BrainQTL (http://predictdb.hakyimlab.org/), local LDSC (https://github.com/bulik/ldsc), NetColoc (https://pypi.org/project/netcoloc/), and Phecode Map 1.2 (https://phewascatalog.org/phecodes).

## ACKNOWLEDGEMENTS

J.J.M., L.V.R., A.A.P., and S.S.R. are funded through the National Institute on Drug Abuse (NIDA DP1DA054394 and P50DA037844). R.B.C., S.R.P., M.V.J., A.A.P., and S.S.R. are funded through the Tobacco-Related Disease Research Program (T32IR5226 and 28IR-0070). H.H.A.T. is supported by a Canadian Institute of Health Research (CIHR) postdoctoral fellowship (#491556). R.B.C is also funded by the Interdisciplinary Research Fellowship in NeuroAIDS (5R25MH081482-17). T.T.M. is supported by funds from the National Institute of Mental Health (K08MH135343). LKD was supported by grants from the National Institutes of Health including R01MH118223.

The PheWAS analysis made use of the VUMC Synthetic Derivative (SD), which was supported by the National Center for Research Resources, Grant UL1 RR024975-01, and is now at the National Center for Advancing Translational Sciences, Grant 2 UL1 TR000445-06. The content is solely the responsibility of the authors and does not necessarily represent the official views of the NIH.

The dataset(s) used for the PheWAS/LabWAS analyses were obtained from Vanderbilt University Medical Center’s BioVU which is supported by numerous sources: institutional funding, private agencies, and federal grants. These include the NIH funded Shared Instrumentation Grant S10RR025141; and CTSA grants UL1TR002243, UL1TR000445, and UL1RR024975. Genomic data are also supported by investigator-led projects that include U01HG004798, R01NS032830, RC2GM092618, P50GM115305, U01HG006378, U19HL065962, R01HD074711; and additional funding sources listed at https://victr.vumc.org/biovu-funding/.

## AUTHOR CONTRIBUTIONS

A.A.P. and S.S.R. conceived the idea and negotiated an agreement with 23andMe that paid for the collection and analysis of the data. P.F. and S.L.E. contributed formal analyses and curation of 23andMe data. S.R.P, P.F., M.V.J., J.J.M, and J.Y. ran analyses. R.B.C, S.R.P., M.V.P, J.J.M, and J.Y. made the figures and tables. All authors reviewed and edited the manuscript.

We would like to thank the research participants and employees of 23andMe for making this work possible. The following members of the 23andMe Research Team contributed to this study:

Stella Aslibekyan, Adam Auton, Elizabeth Babalola, Robert K. Bell, Jessica Bielenberg, Ninad S. Chaudhary, Zayn Cochinwala, Sayantan Das, Emily DelloRusso, Payam Dibaeinia, Sarah L. Elson, Nicholas Eriksson, Chris Eijsbouts, Teresa Filshtein, Pierre Fontanillas, Davide Foletti, Will Freyman, Zach Fuller, Julie M. Granka, Chris German, Éadaoin Harney, Alejandro Hernandez, Barry Hicks, David A. Hinds, M. Reza Jabalameli, Ethan M. Jewett, Yunxuan Jiang, Sotiris Karagounis, Lucy Kaufmann, Matt Kmiecik, Katelyn Kukar, Alan Kwong, Keng-Han Lin, Yanyu Liang, Bianca A. Llamas, Aly Khan, Steven J. Micheletti, Matthew H. McIntyre, Meghan E. Moreno, Priyanka Nandakumar, Dominique T. Nguyen, Jared O’Connell, Steve Pitts, G. David Poznik, Alexandra Reynoso, Shubham Saini, Morgan Schumacher, Leah Selcer, Anjali J. Shastri, Jingchunzi Shi, Suyash Shringarpure, Keaton Stagaman, Teague Sterling, Qiaojuan Jane Su, Joyce Y. Tung, Susana A. Tat, Vinh Tran, Xin Wang, Wei Wang, Catherine H. Weldon, Amy L. Williams, Peter Wilton.

## COMPETING INTERESTS

P.F., the 23andMe Research Team, and S.L.E. are employed by 23andMe, Inc. P.F. and S.L.E. hold stock or stock options in 23andMe, Inc. The remaining authors have nothing to disclose.

