## Supplementary Material for "Genome-wide association study of delay discounting in 134,935 individuals identifies novel loci and transdiagnostic associations across mental and physical health"

### **Supplementary Methods**

#### Monetary Choice Questionnaire

Over the course of approximately 4 months in 2015, over 134,000 individuals responded to survey questions as part of a study on the genetics of decision-making developed by A.A.P. and S.S.R. This included a 30-item monetary choice questionnaire (**MCQ**) we previously employed^1^, which was modified from the well-established 27-item MCQ^2^. The original 27 items presented individuals with a choice between a smaller immediate reward and a larger delayed reward at one of three magnitudes (small: $25–35; intermediate: $50–60; large: $75–85). Additionally, 3 items were introduced to the original 27-item MCQ specifically aimed at excluding inappropriate responders. The questions were as follows:

| Item | Would you rather have: | |
| --- | --- | --- |
| 1 | $54 Today | $55 in 117 Days |
| 2 | $55 Today | $75 in 61 Days |
| 3 | $19 Today | $25 in 53 Days |
| 4 | $31 Today | $85 in 7 Days |
| 5 | $14 Today | $25 in 19 Days |
| 6 | $47 Today | $50 in 160 Days |
| 7 | $15 Today | $35 in 13 Days |
| 8* | $55 Today | $85 Today |
| 9 | $25 Today | $60 in 14 Days |
| 10 | $78 Today | $80 in 162 Days |
| 11 | $40 Today | $55 in 62 Days |
| 12 | $11 Today | $30 in 7 Days |
| 13 | $67 Today | $75 in 119 Days |
| 14 | $34 Today | $35 in 186 Days |
| 15 | $27 Today | $50 in 21 Days |
| 16 | $69 Today | $85 in 91 Days |
| 17* | $60 Today | $20 Today |
| 18 | $49 Today | $60 in 89 Days |
| 19 | $80 Today | $85 in 157 Days |
| 20 | $24 Today | $35 in 29 Days |
| 21 | $33 Today | $80 in 14 Days |
| 22 | $28 Today | $30 in 179 Days |
| 23 | $34 Today | $50 in 30 Days |
| 24* | $15 Today | $35 Today |
| 25 | $25 Today | $30 in 80 Days |
| 26 | $41 Today | $75 in 20 Days |
| 27 | $54 Today | $60 in 111 Days |
| 28 | $54 Today | $80 in 30 Days |
| 29 | $22 Today | $25 in 136 Days |
| 30 | $20 Today | $55 in 7 Days |

*These items were not part of the original MCQ. We added them to identify individuals who responded carelessly. Participants choosing one or more lower monetary rewards for items 8, 17 or 24 were presumed to be responding carelessly and excluded from all analysis.

The overall response pattern was used to derive temporal discounting functions (***k***) spanning 1.60×10^-04^ to 0.25. Higher values of k indicate a greater devaluation of delayed rewards, reflecting a stronger preference for immediate gratification. In the full cohort, most research participants (~98%) demonstrated high *k* consistency across questions. Participants with less than 80% concordance across the three reward magnitudes were excluded from the analyses. The inappropriate response rate was 1.70% (*N* = 3,230) in the full cohort. *k* values were not normally distributed; we employed a log-10 transformation, a common technique used to approximate a normal distribution for *k*^3^. The log10(*k*) values were averaged over the three reward magnitudes.

#### Analysis of local ancestry

We performed delay discounting (DD) genome-wide association study (GWAS) on 134,935 23andMe, Inc. participants classified as being of European genetic similarity. Ancestry falls along a spectrum; each individual was clustered based on genetic similarity to a reference panel using local ancestry analysis. Briefly, the 23andMe algorithm first partitions phased genomic data into short windows of about 300 SNPs. Within each window, we use a support vector machine to classify individual haplotypes into one of 31 reference populations (<https://www.23andme.com/ancestry-composition-guide/>). The support vector machine classifications are fed into a hidden Markov model that accounts for switch errors and incorrect assignments, and gives probabilities for each reference population in each window. The reference population data is derived from public datasets (the Human Genome Diversity Project, HapMap, and 1000 Genomes), as well as 23andMe customers who have reported having four grandparents from the same country.

Genetic similarity is defined as follows:

| **Ancestry** | **Classification Criteria** |
| --- | --- |
| European | European + Middle Eastern > 0.97, European > 0.90 |
| East Asian | East Asian + Southeast Asian > 0.97 |
| South Asian | South Asian > 0.97 |
| Middle Eastern (& North African) | Middle Eastern + European > 0.97, Middle Eastern > 0.90 |
| African American + Latin American | European + African + East Asian + Native American + Middle Eastern > 0.90, African + Native American > 0.01 |

#### Genotyping, relatedness, and imputation

Sample genotyping was performed on a custom genotyping array platform developed by 23andMe, which includes the Illumina HumanHap550+ Bead chip V1 V2, OmniExpress+ Bead chip V3, and custom array V4. All samples met a minimum call rate of 98.5%. In total, 1,609,130 single SNPs and insertions/deletions were genotyped across all platforms (for further details on genotyping and sample quality, see^4–6^). We phased and imputed data for each genotyping platform separately.

A comprehensive set of unrelated individuals was selected using a segmental identity-by-descent (**IBD**) estimation algorithm^7^ to ensure that only unrelated individuals were included in the sample. Individuals were classified as related if they shared more than 700cM IBD, including regions where the two individuals shared either one or both genomic segments IBD. This level of relatedness (~20% of the genome) corresponds to approximately the minimal expected sharing between first cousins in an outbred population.

Imputation of genotype data was based on the 1,000 Genomes phase 1 version 3 reference haplotypes. The data were phased and imputed separately for each genotyping platform. Phasing was performed using an in-house phasing tool called Finch, which implements the Beagle haplotype graph-based phasing algorithm^8^ that is modified to separate the haplotype graph construction and phasing steps. To prepare for imputation, we split phased chromosomes into segments containing no more than 10,000 genotyped SNPs, with overlaps of 200 SNPs. SNPs were excluded if they had Hardy-Weinberg equilibrium *P* < 1.00×10^−20^, call rate < 90%, or exhibited large allele frequency discrepancies compared to the European 1,000 Genomes reference data. These discrepancies were identified by computing a 2 × 2 table of allele counts for European 1,000 Genomes samples and 2,000 randomly sampled 23andMe customers with European genetic similarity. SNPs with *P* < 1.00×10^−15^ by *χ^2^* test were flagged as having significant frequency discrepancies. Each phased segment was imputed against all-ancestry 1,000 Genomes haplotypes (excluding monomorphic and singleton sites) using Minimac2^9^, with five rounds and 200 states for parameter estimation. Following imputation and quality control procedures, we analyzed a total of 14,137,232 SNPs.

Regarding the X chromosome, distinct haplotype graphs were constructed for both the non-pseudoautosomal region and each pseudoautosomal region. Phasing was subsequently performed separately for these regions. For imputation, both males and females were imputed together using Minimac2, following the same procedure as with the autosomes. In the non-pseudoautosomal region, males were treated as homozygous pseudodiploids.

For analyses involving imputed data, we utilized imputed dosages rather than the best-guess genotypes. HLA allele dosages were imputed from SNP genotype data using HIBAG^10^. Imputation was conducted for alleles at the HLA-A, -B, -C, DPB1, DQA1, DQB1, and DRB1 loci at a four-digit resolution. To examine associations between HLA allele dosages and phenotypes, linear regression was performed, employing the same set of covariates utilized in the SNP-based GWAS (i.e., age, 5 principal components and genotype platform). Separate association tests were conducted for each imputed allele. HLA did not reveal any significant signal and thus is not elaborated.

For GWAS analysis, SNPs were excluded if they had a minor allele frequency < 0.1%, failed Hardy–Weinberg equilibrium (*P* < 1.00×10^−20^), had a call rate < 90%, failed a Mendelian transmission test in trios (*P* < 1.00×10^−20^), showed large allele frequency discrepancies compared to European 1000 Genomes reference data, or failed batch effects testing (ANOVA *P* < 1.00×10^−20^). Imputed variants were excluded if they showed low imputation quality (*r^2^* < 0.50 averaged across batches or minimum *r^2^* < 0.30) or showed evidence of batch effects (ANOVA *P* < 1.00×10^−50^). The top 5 principal components of genotype that were used as model covariates were computed using ~65,000 high-quality genotyped variants present in all four genotyping platforms.

#### Credible set analysis

#### This credible set is calculated by first calculating Wakefield’s Approximate Bayes Factor (ABFs) for each SNP and assuming a prior variance (W) of 0.1. The credible set is then estimated from the ABFs using the method of Maller et al^11^.

#### Gene- and transcriptome-based analysis

To identify independent SNPs and their functional consequences, we utilized the FUMA (v1.3.6a) web-based platform^12^. Independent SNPs were defined as those with a linkage disequilibrium coefficient *r^2^* < 0.10. We used Multi-marker Analysis of GenoMic Annotation (**MAGMA**; v1.08)^13^ to perform gene-set analysis. SNPs were mapped to 19,728 protein-coding genes sourced from Ensembl (build 85). We applied a Bonferroni threshold on the total number of genes tested (*P* < 2.53×10^-6^).

We used Hi-C coupled MAGMA (**H-MAGMA**)^14,15^ to assign intergenic and intronic SNPs to genes based on their chromatin interactions. Exonic and promoter SNPs were assigned to genes according to their physical position. H-MAGMA integrates data from four Hi-C datasets derived from fetal brain, adult brain, iPSC-derived neurons, and iPSC-derived astrocytes^16^. We applied a Bonferroni correction based on the total number of gene-tissue pairs tested (*P* < 9.78×10^-7^).

We used S-PrediXcan to incorporate genomic results with transcriptomic data and identify specific expression quantitative trait loci-linked genes associated with DD. This method leverages pre-computed tissue weights from the Genotype-Tissue Expression (GTEx) v8 project database^17–19^ to predict transcript levels and evaluates whether these predicted transcripts correlate with DD using a sparse (elastic net) prediction model. We conducted this analysis in 13 brain tissues available through GTEx. We applied a Bonferroni threshold based on the number of genes (*N* = 2,532-6,744; *P* < 1.97×10^-5^ to 7.41×10^-6^).

### **Supplementary Figures**

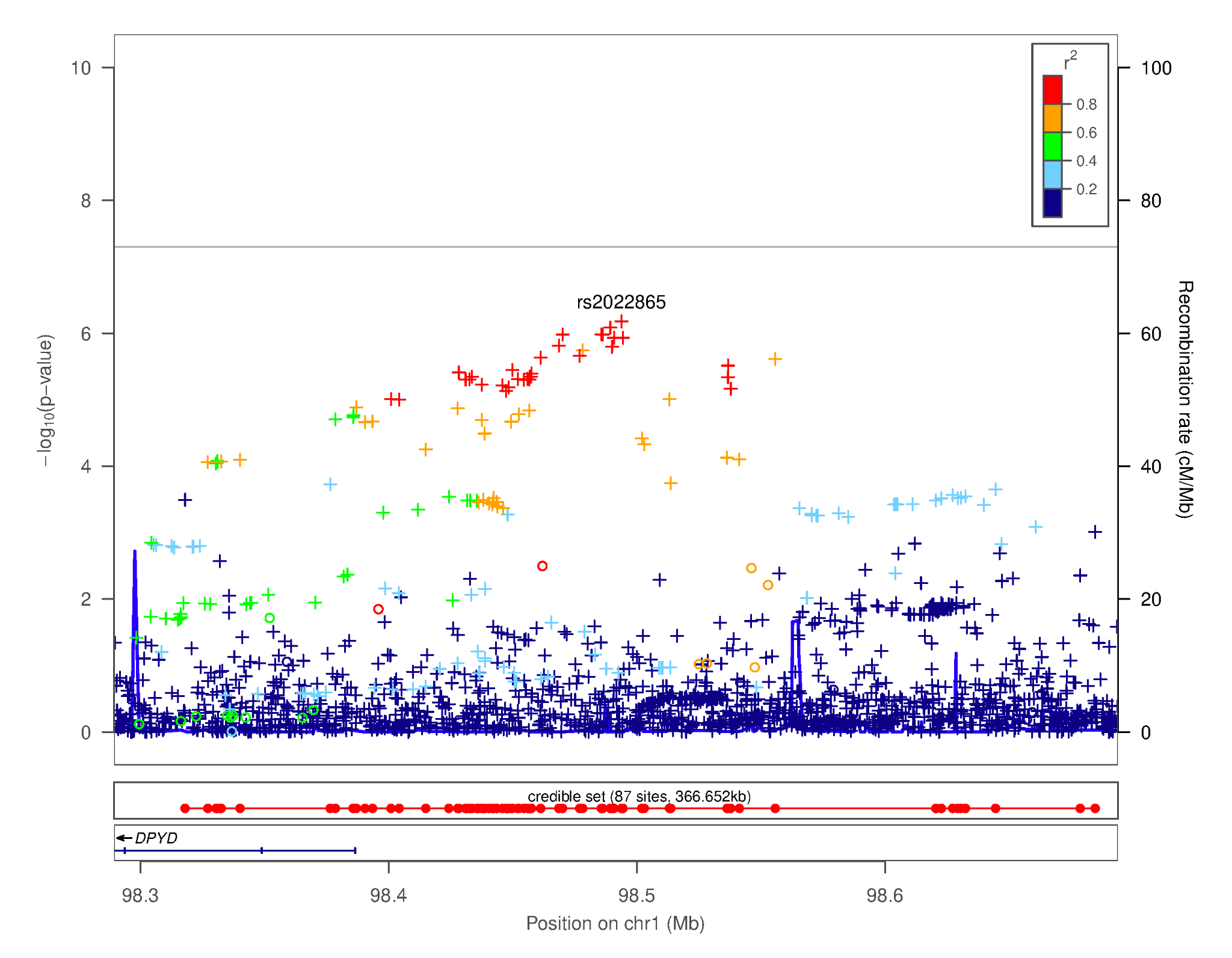

#### Supplementary Figure 1. Regional association plot focusing on the region chr1p21.3 - most significant SNP: rs2022865 [*DPYD*—[]—*SNX7*]. This plot was generated using LocusZoom^20^. The -log_10_(*p-*value) is shown on the left *y-*axis; position in Mb is on the *x*-axis. Recombination rates (expressed in centiMorgans cM per Mb; NCBI Build GRCh37; highlighted in blue) are shown on the right *y-*axis. Pairwise linkage disequilibrium (*r^2^*) of each SNP with the top SNP in the region is indicated by its color. Crossed points represent imputed SNPs, circles represent directly genotyped SNPs.

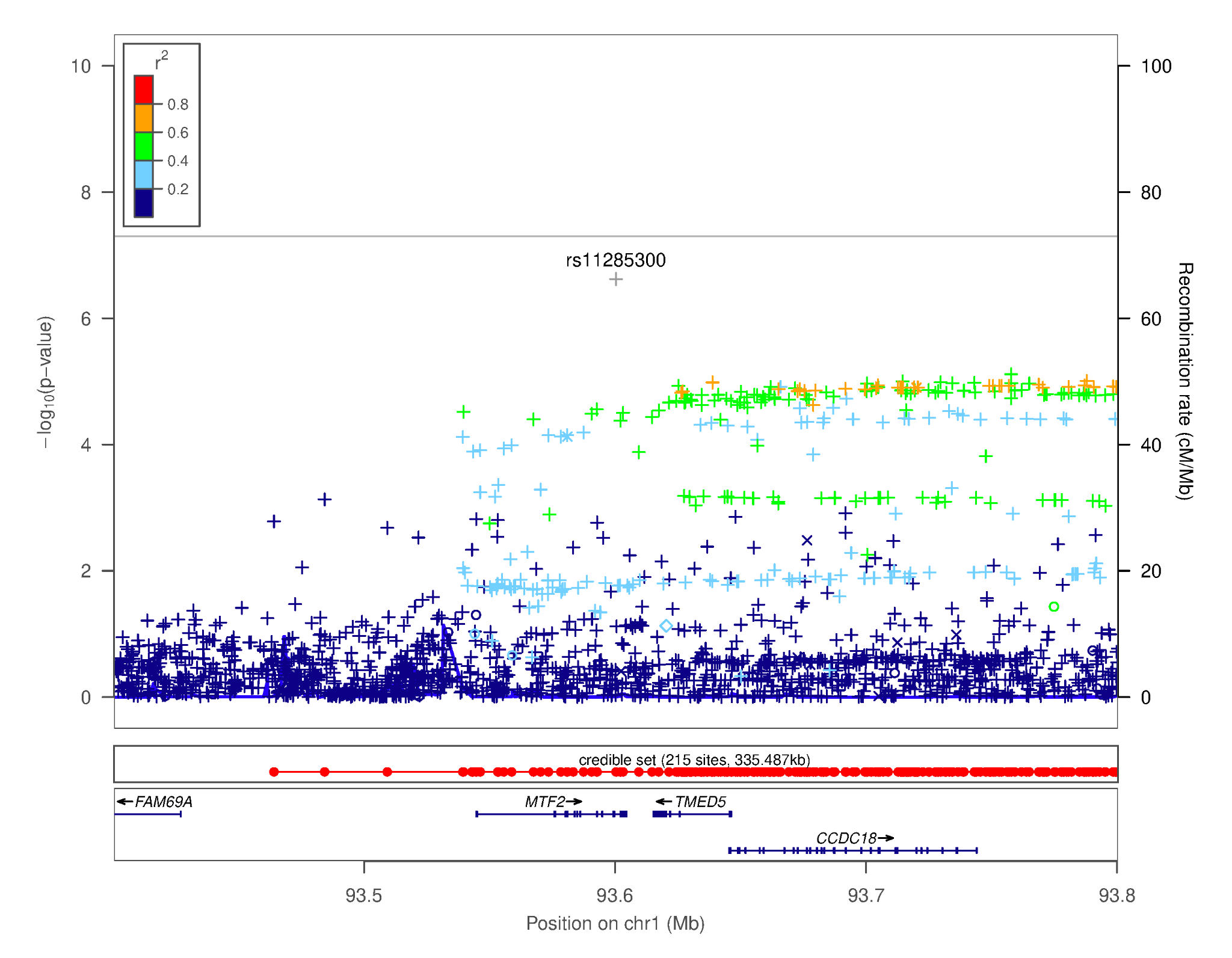

#### Supplementary Figure 2. Regional association plot focusing on the region chr1p22.1- most significant SNP: rs11285300 [*MTF2*]. This plot was generated using LocusZoom^20^. The -log_10_(*p-*value) is shown on the left *y-*axis; position in Mb is on the *x*-axis. Recombination rates (expressed in centiMorgans cM per Mb; NCBI Build GRCh37; highlighted in blue) are shown on the right *y-*axis. Pairwise linkage disequilibrium (*r^2^*) of each SNP with the top SNP in the region is indicated by its color. Crossed points represent imputed SNPs, circles represent directly genotyped SNPs.

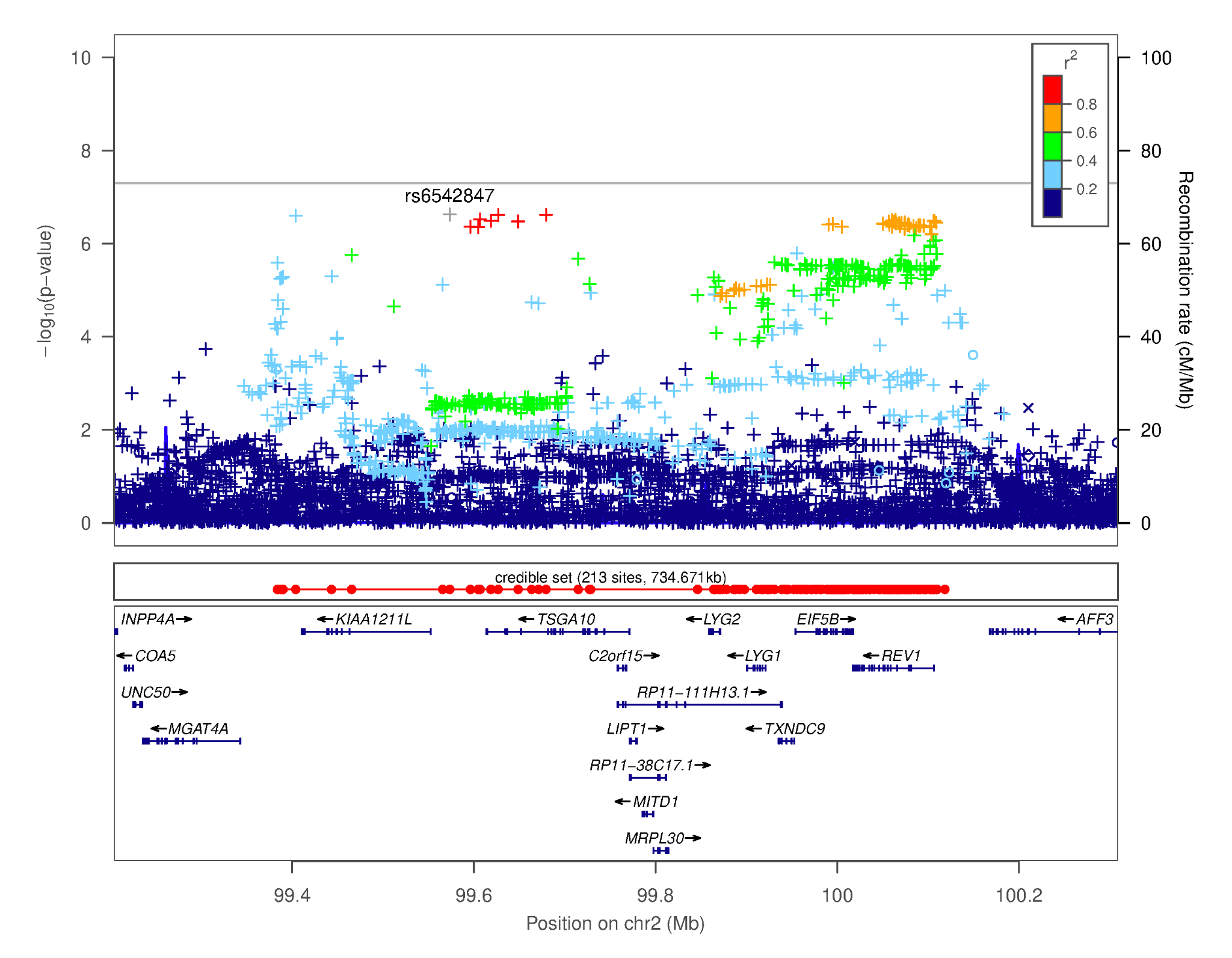

#### Supplementary Figure 3. Regional association plot focusing on the region chr2q11.2- most significant SNP: rs6542847 [*KIAA1211L*–[]*–TSGA10*]. This plot was generated using LocusZoom^20^. The -log_10_(*p-*value) is shown on the left *y-*axis; position in Mb is on the *x*-axis. Recombination rates (expressed in centiMorgans cM per Mb; NCBI Build GRCh37; highlighted in blue) are shown on the right *y-*axis. Pairwise linkage disequilibrium (*r^2^*) of each SNP with the top SNP in the region is indicated by its color. Crossed points represent imputed SNPs, circles represent directly genotyped SNPs.

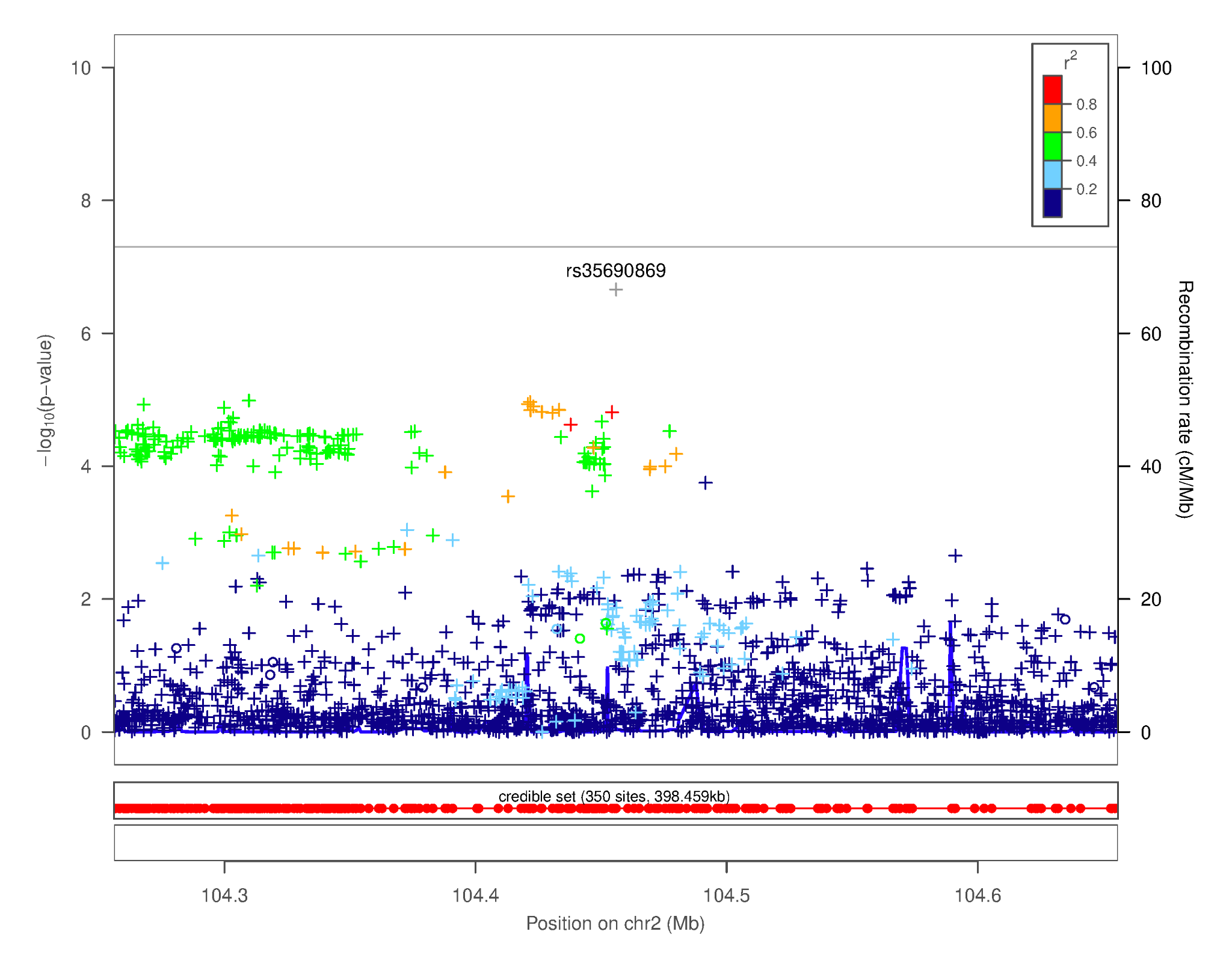

#### Supplementary Figure 4. Regional association plot focusing on the region chr2q12.1- most significant SNP: rs35690869. This plot was generated using LocusZoom^20^. The -log_10_(*p-*value) is shown on the left *y-*axis; position in Mb is on the *x*-axis. Recombination rates (expressed in centiMorgans cM per Mb; NCBI Build GRCh37; highlighted in blue) are shown on the right *y-*axis. Pairwise linkage disequilibrium (*r^2^*) of each SNP with the top SNP in the region is indicated by its color. Crossed points represent imputed SNPs, circles represent directly genotyped SNPs.

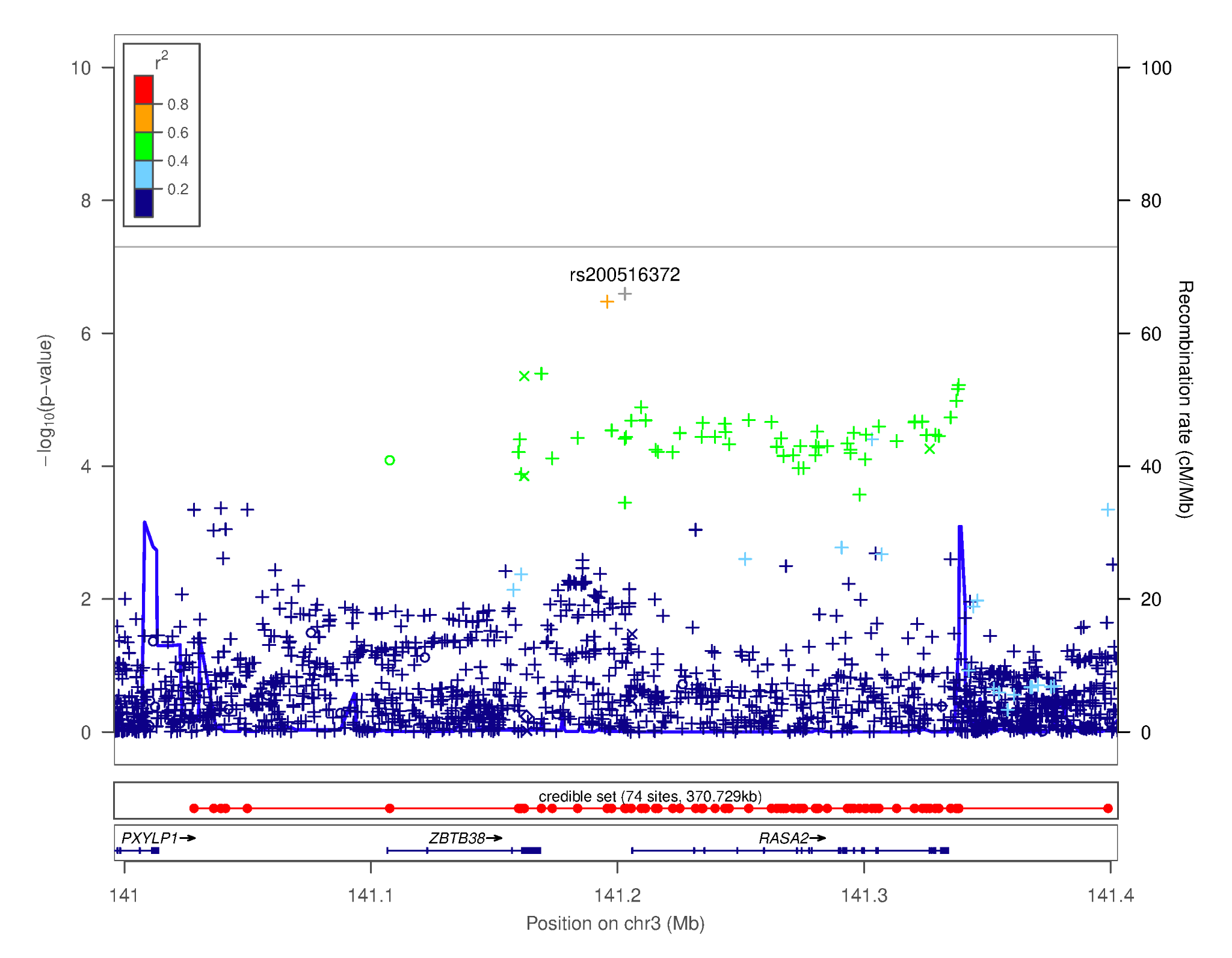

#### Supplementary Figure 5. Regional association plot focusing on the region chr3q23- most significant SNP: rs200516372 [*ZBTB38–*[]-*RASA2*]. This plot was generated using LocusZoom^20^. The -log_10_(*p-*value) is shown on the left *y-*axis; position in Mb is on the *x*-axis. Recombination rates (expressed in centiMorgans cM per Mb; NCBI Build GRCh37; highlighted in blue) are shown on the right *y-*axis. Pairwise linkage disequilibrium (*r^2^*) of each SNP with the top SNP in the region is indicated by its color. Crossed points represent imputed SNPs, circles represent directly genotyped SNPs.

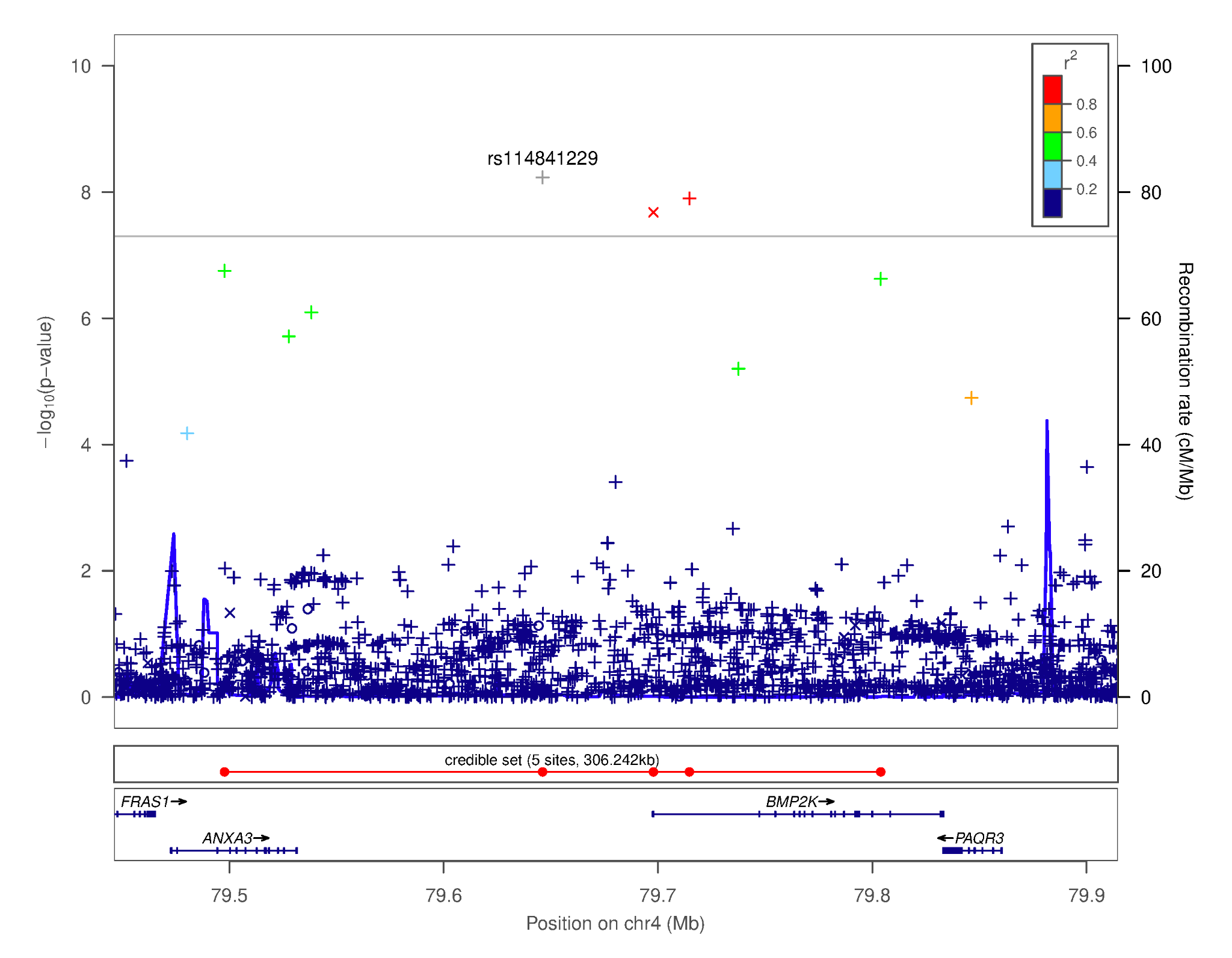

#### Supplementary Figure 6. Regional association plot focusing on the region chr4q21.21- most significant SNP: rs114841229 [*ANXA3*—[]–*BMP2K*]. This plot was generated using LocusZoom^20^. The -log_10_(*p-*value) is shown on the left *y-*axis; position in Mb is on the *x*-axis. Recombination rates (expressed in centiMorgans cM per Mb; NCBI Build GRCh37; highlighted in blue) are shown on the right *y-*axis. Pairwise linkage disequilibrium (*r^2^*) of each SNP with the top SNP in the region is indicated by its color. Crossed points represent imputed SNPs, circles represent directly genotyped SNPs.

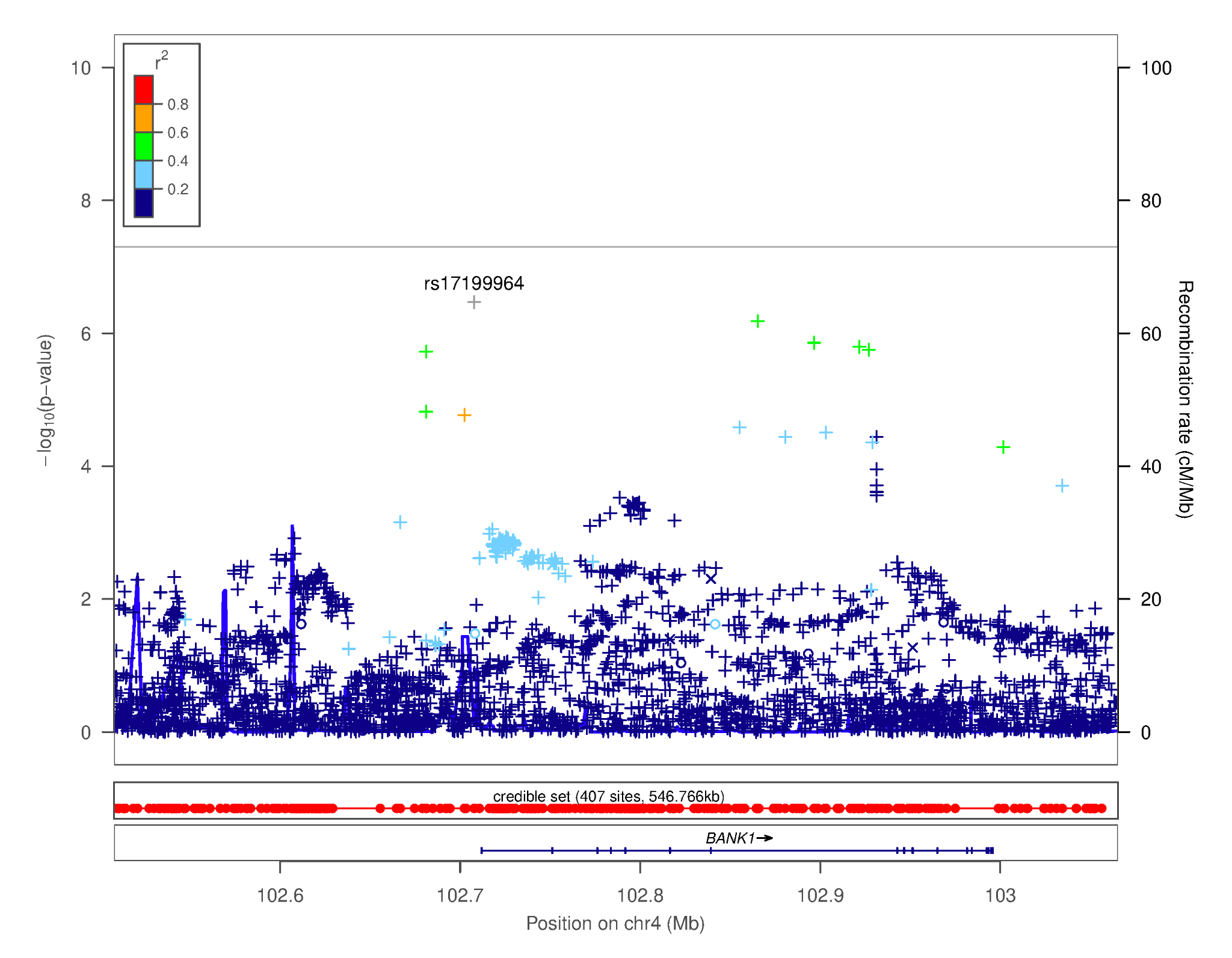

#### Supplementary Figure 7. Regional association plot focusing on the region chr4q24- most significant SNP: rs17199964 [*PPP3CA*—[]-*BANK1*]. This plot was generated using LocusZoom^20^. The -log_10_(*p-*value) is shown on the left *y-*axis; position in Mb is on the *x*-axis. Recombination rates (expressed in centiMorgans cM per Mb; NCBI Build GRCh37; highlighted in blue) are shown on the right *y-*axis. Pairwise linkage disequilibrium (*r^2^*) of each SNP with the top SNP in the region is indicated by its color. Crossed points represent imputed SNPs, circles represent directly genotyped SNPs.

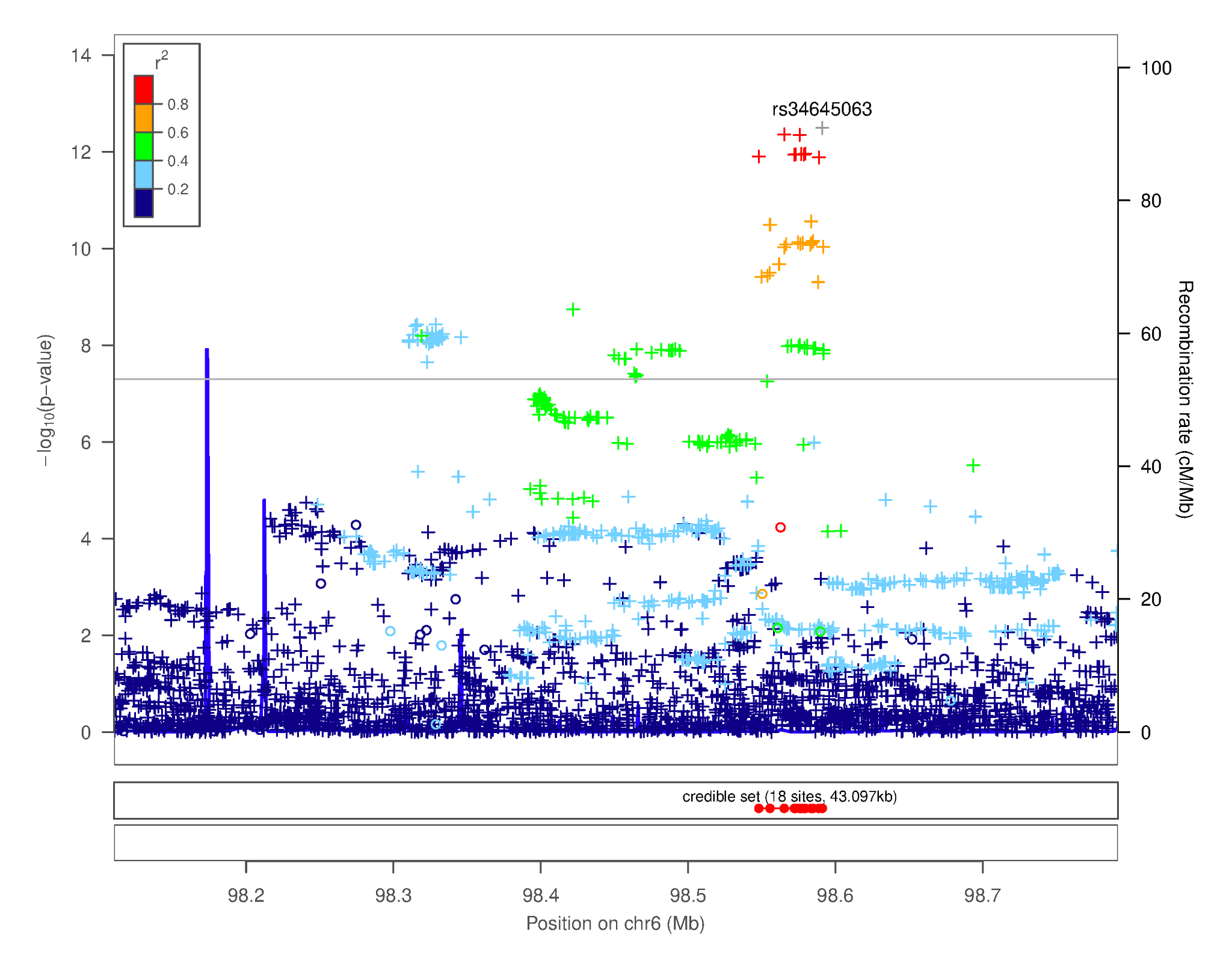

#### Supplementary Figure 8. Regional association plot focusing on the region chr6q16.1- most significant SNP: rs34645063 [*MMS22L—*[]—*POU3F2*]. This plot was generated using LocusZoom^20^. The -log_10_(*p-*value) is shown on the left *y-*axis; position in Mb is on the *x*-axis. Recombination rates (expressed in centiMorgans cM per Mb; NCBI Build GRCh37; highlighted in blue) are shown on the right *y-*axis. Pairwise linkage disequilibrium (*r^2^*) of each SNP with the top SNP in the region is indicated by its color. Crossed points represent imputed SNPs, circles represent directly genotyped SNPs.

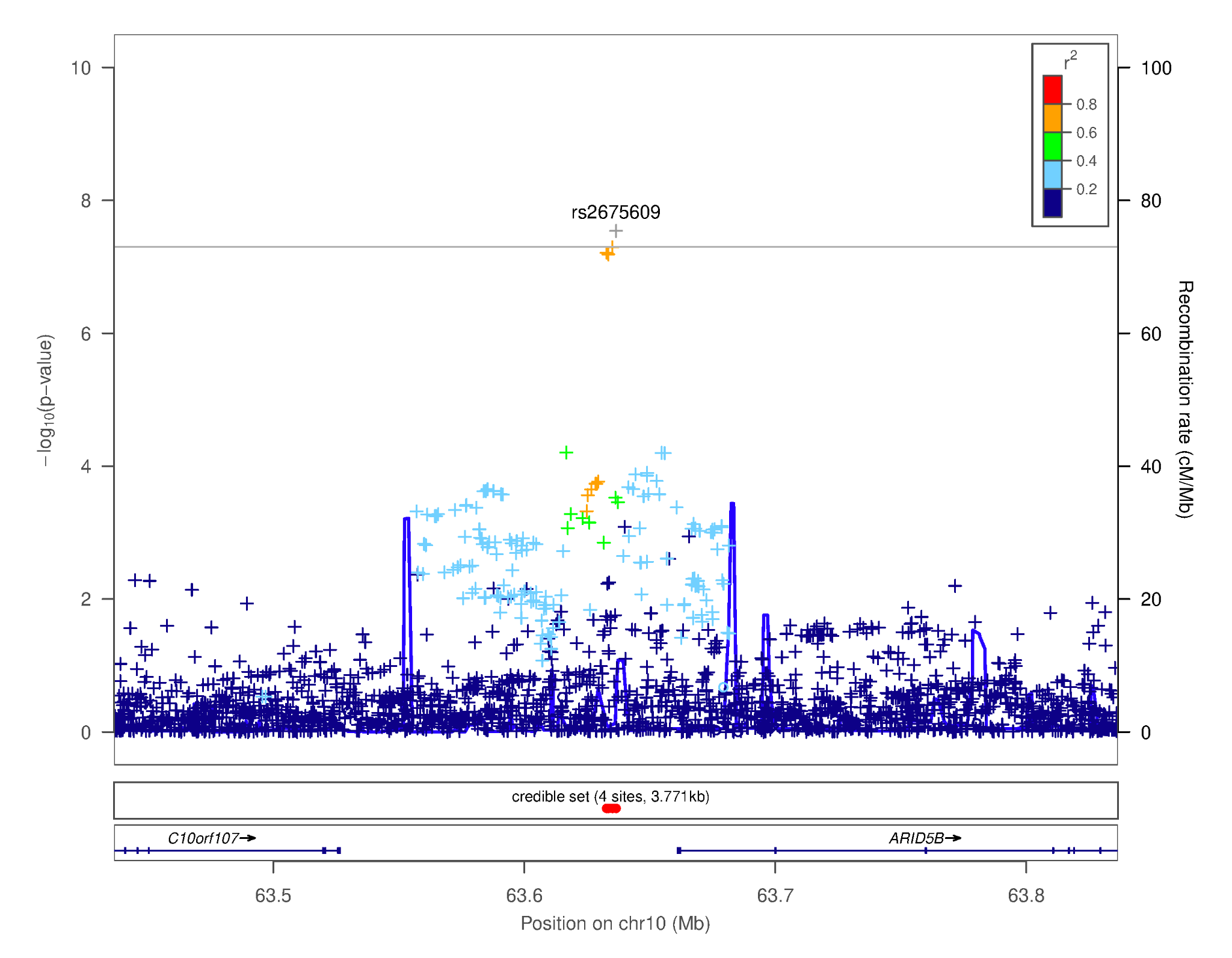

#### Supplementary Figure 9. Regional association plot focusing on the region chr10q21.2- most significant SNP: rs200581586 [*RP11-298I3.5*]. This plot was generated using LocusZoom^20^. The -log_10_(*p-*value) is shown on the left *y-*axis; position in Mb is on the *x*-axis. Recombination rates (expressed in centiMorgans cM per Mb; NCBI Build GRCh37; highlighted in blue) are shown on the right *y-*axis. Pairwise linkage disequilibrium (*r^2^*) of each SNP with the top SNP in the region is indicated by its color. Crossed points represent imputed SNPs, circles represent directly genotyped SNPs.

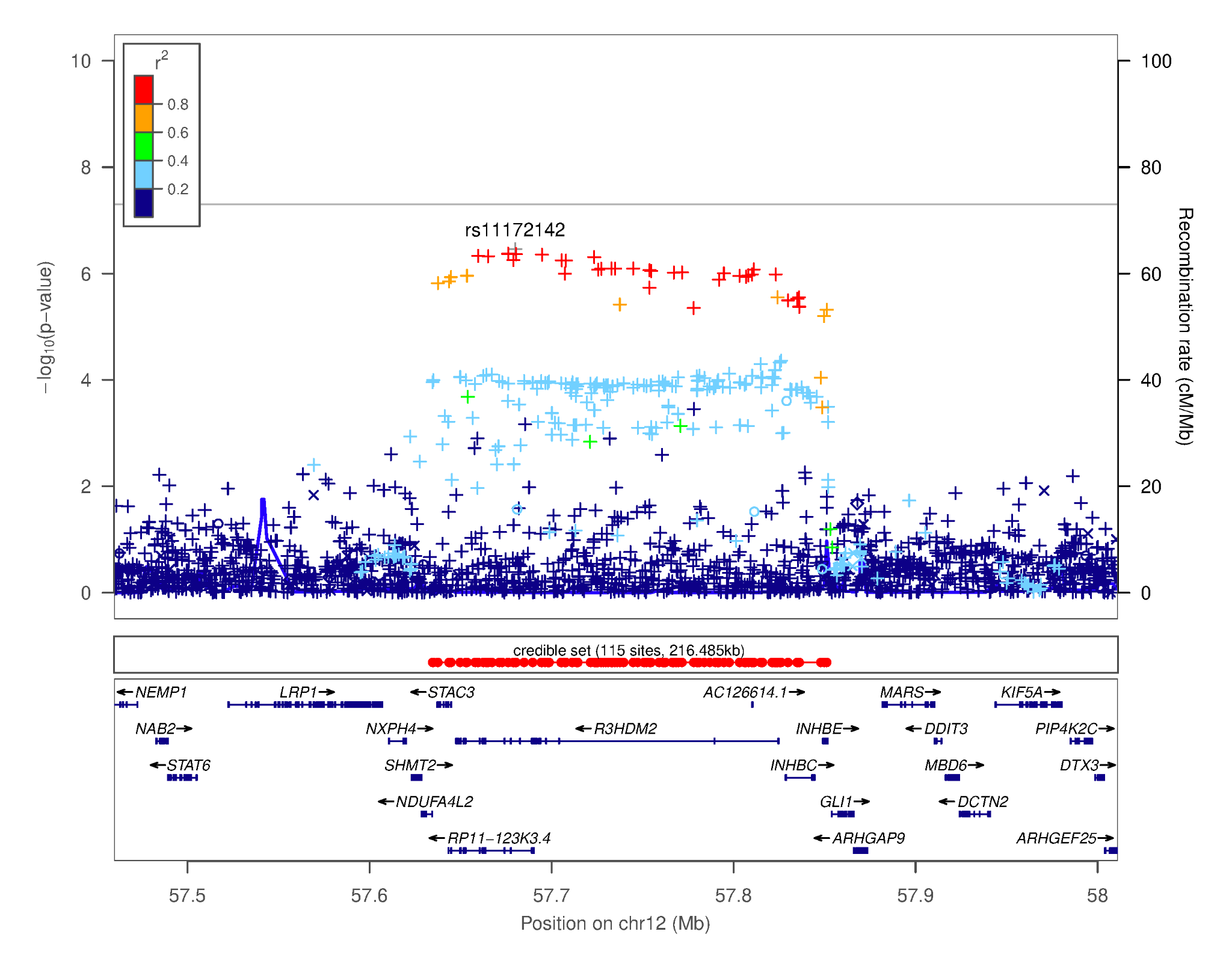

#### Supplementary Figure 10. Regional association plot focusing on the region chr12q13.3- most significant SNP: rs11172142 [*RP11-123K3.4,R3HDM2*]. This plot was generated using LocusZoom^20^. The -log_10_(*p-*value) is shown on the left *y-*axis; position in Mb is on the *x*-axis. Recombination rates (expressed in centiMorgans cM per Mb; NCBI Build GRCh37; highlighted in blue) are shown on the right *y-*axis. Pairwise linkage disequilibrium (*r^2^*) of each SNP with the top SNP in the region is indicated by its color. Crossed points represent imputed SNPs, circles represent directly genotyped SNPs.

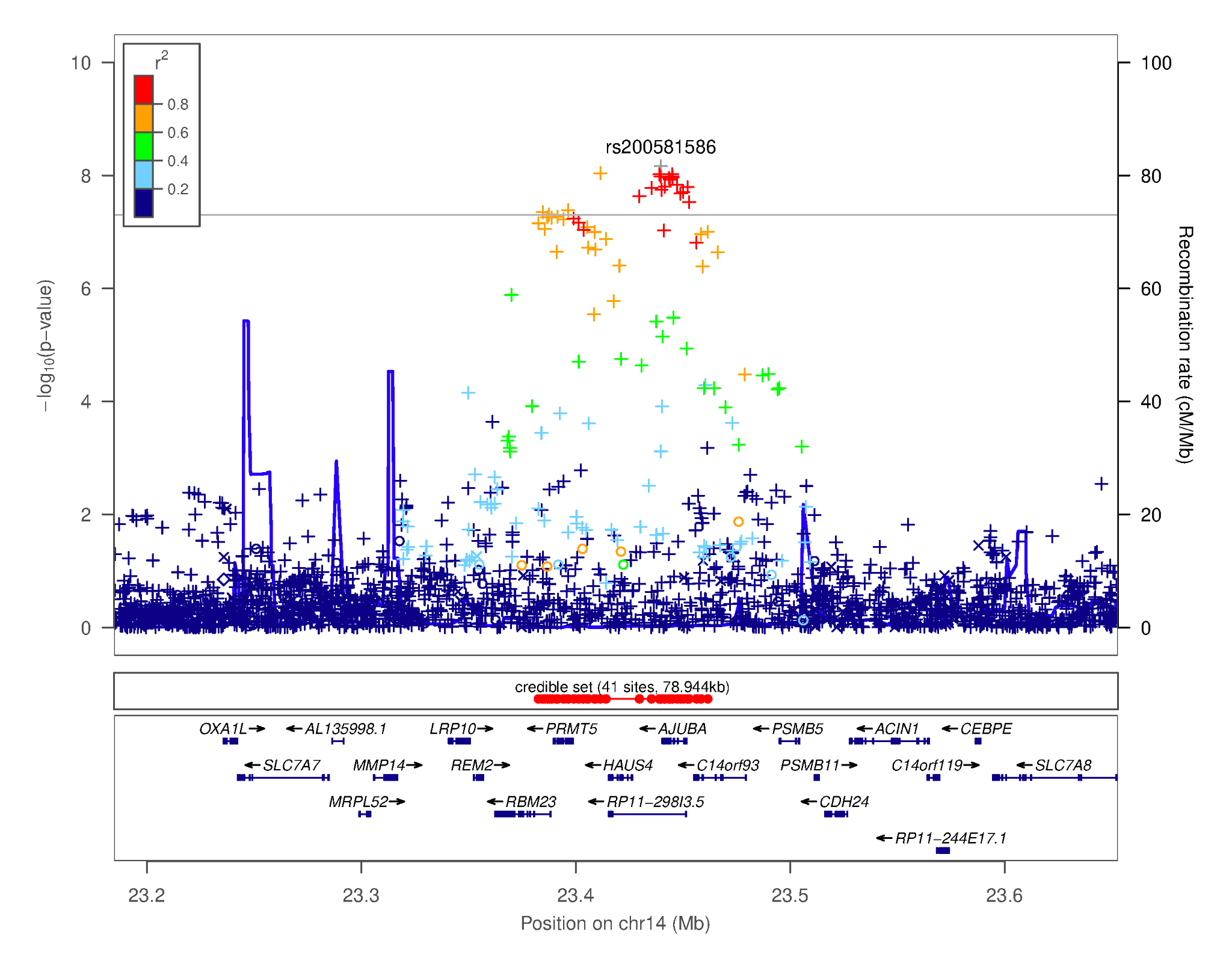

#### Supplementary Figure 11. Regional association plot focusing on the region chr14q11.2- most significant SNP: rs200581586 [*RP11-298I3.5*]. This plot was generated using LocusZoom^20^. The -log_10_(*p-*value) is shown on the left *y-*axis; position in Mb is on the *x*-axis. Recombination rates (expressed in centiMorgans cM per Mb; NCBI Build GRCh37; highlighted in blue) are shown on the right *y-*axis. Pairwise linkage disequilibrium (*r^2^*) of each SNP with the top SNP in the region is indicated by its color. Crossed points represent imputed SNPs, circles represent directly genotyped SNPs.

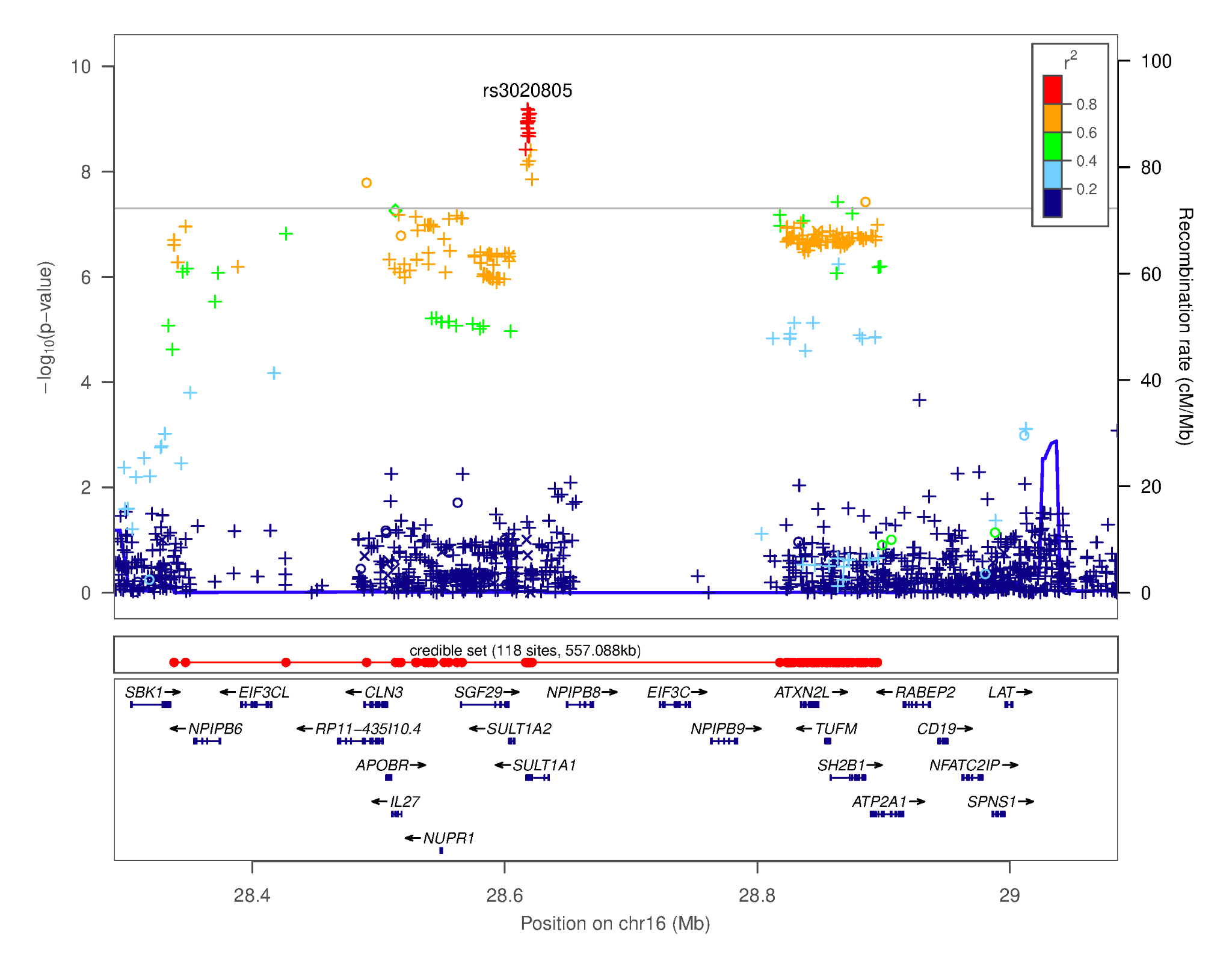

#### Supplementary Figure 12. Regional association plot focusing on the region chr16p11.2- most significant SNP: rs3020805 [*SULT1A1*]. This plot was generated using LocusZoom^20^. The -log_10_(*p-*value) is shown on the left *y-*axis; position in Mb is on the *x*-axis. Recombination rates (expressed in centiMorgans cM per Mb; NCBI Build GRCh37; highlighted in blue) are shown on the right *y-*axis. Pairwise linkage disequilibrium (*r^2^*) of each SNP with the top SNP in the region is indicated by its color. Crossed points represent imputed SNPs, circles represent directly genotyped SNPs.

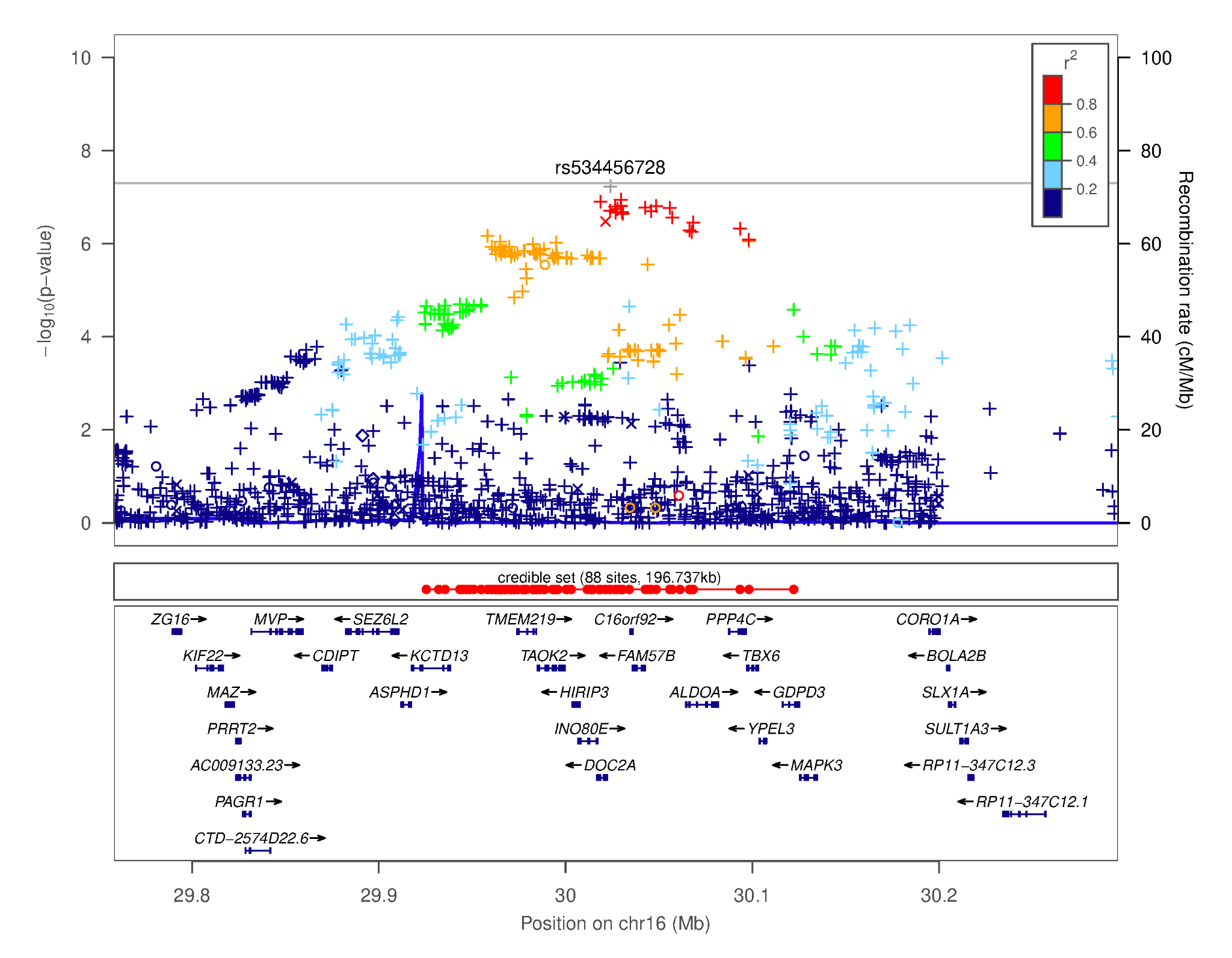

#### Supplementary Figure 13. Regional association plot focusing on the region chr16p11.2- most significant SNP: rs534456728 [*DOC2A*-[]–*C16orf92*]. This plot was generated using LocusZoom^20^. The -log_10_(*p-*value) is shown on the left *y-*axis; position in Mb is on the *x*-axis. Recombination rates (expressed in centiMorgans cM per Mb; NCBI Build GRCh37; highlighted in blue) are shown on the right *y-*axis. Pairwise linkage disequilibrium (*r^2^*) of each SNP with the top SNP in the region is indicated by its color. Crossed points represent imputed SNPs, circles represent directly genotyped SNPs.

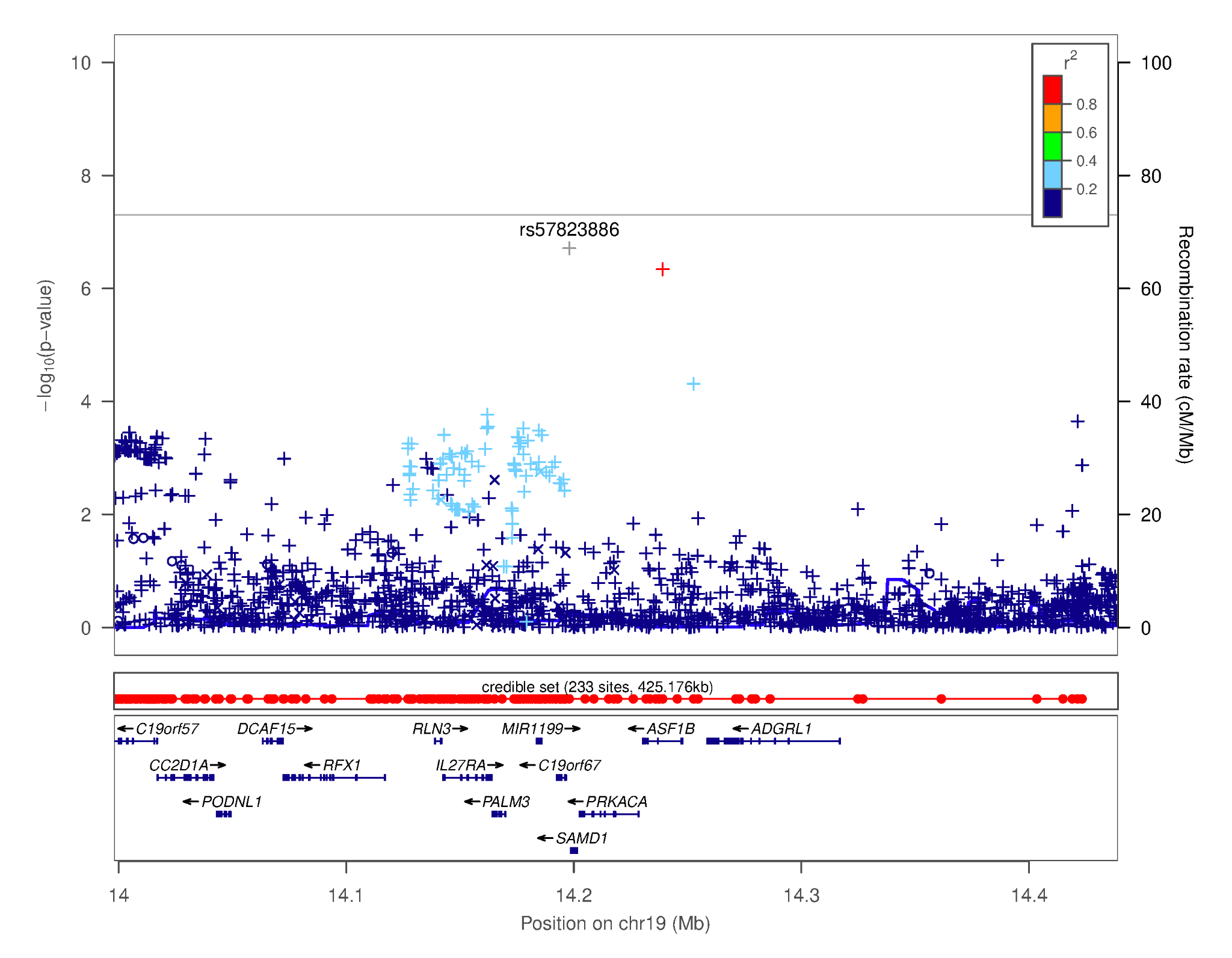

#### Supplementary Figure 14. Regional association plot focusing on the region chr19p13.12- most significant SNP: rs57823886 [*C19orf67*-[]*SAMD1*]. This plot was generated using LocusZoom^20^. The -log_10_(*p-*value) is shown on the left *y-*axis; position in Mb is on the *x*-axis. Recombination rates (expressed in centiMorgans cM per Mb; NCBI Build GRCh37; highlighted in blue) are shown on the right *y-*axis. Pairwise linkage disequilibrium (*r^2^*) of each SNP with the top SNP in the region is indicated by its color. Crossed points represent imputed SNPs, circles represent directly genotyped SNPs.

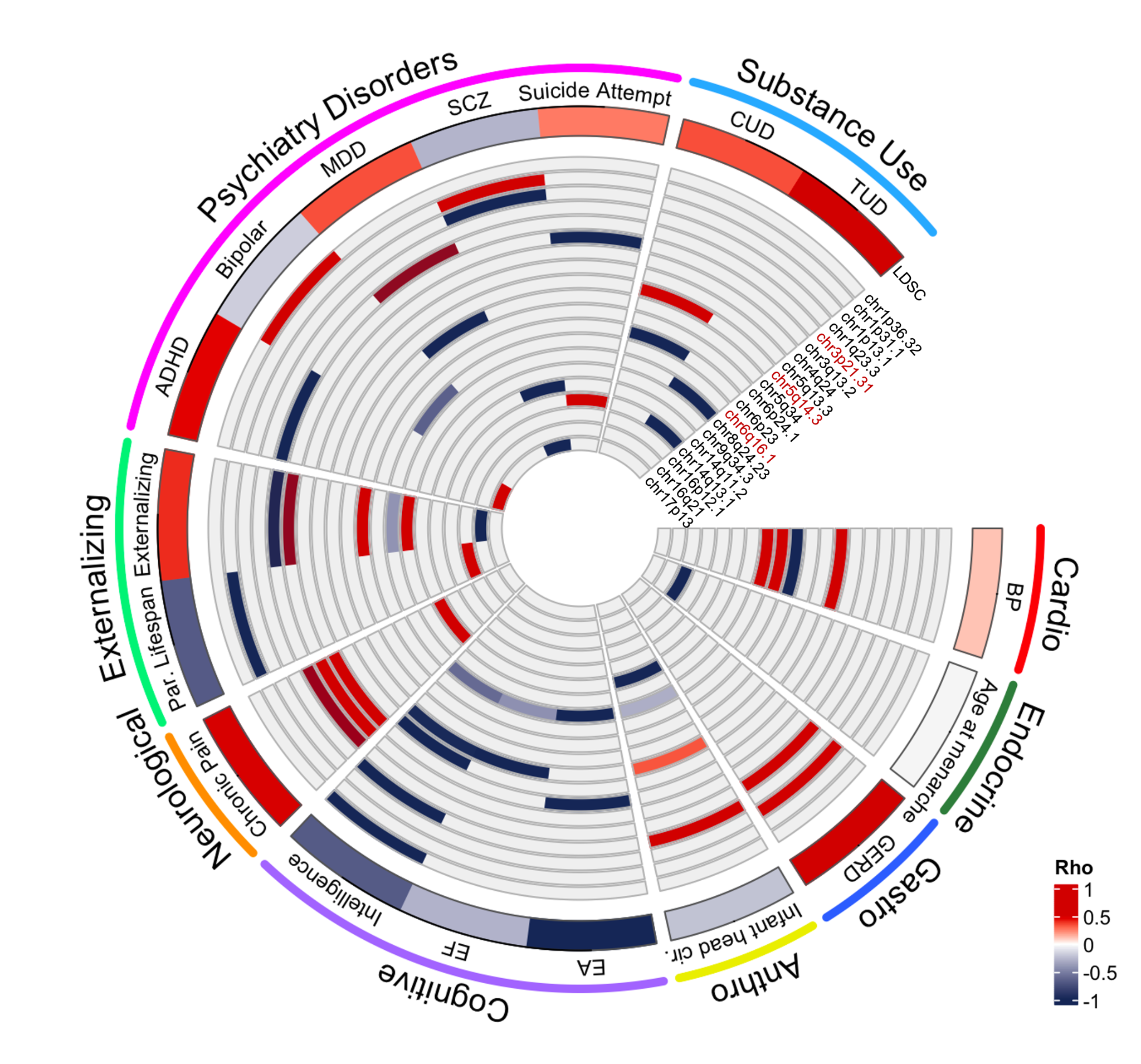

#### Supplementary Figure 15. Plot depicting local genetic correlations across trait categories for loci with 2 or more FDR-significant local genetic correlations (inner rings) with DD. LDSC global genetic correlations for each displayed trait shown in the outer ring. Abbreviations: Cardiovascular (Cardio); Gastrointestinal (Gastro); Infant head circumference at 6-30 months old (Infant head cir.); Anthropometric (Anthro); Blood Pressure (BP); Gastroesophageal Reflux Disease (GERD); Educational attainment (EA); Executive function (EF); Parental lifespan (Par. Lifespan); Attention-deficit hyperactivity disorder (ADHD); Major depressive disorder (MDD); Schizophrenia (SCZ); Cannabis use disorder (CUD); Tobacco use disorder (TUD). For full results, see Supplementary Table 13.

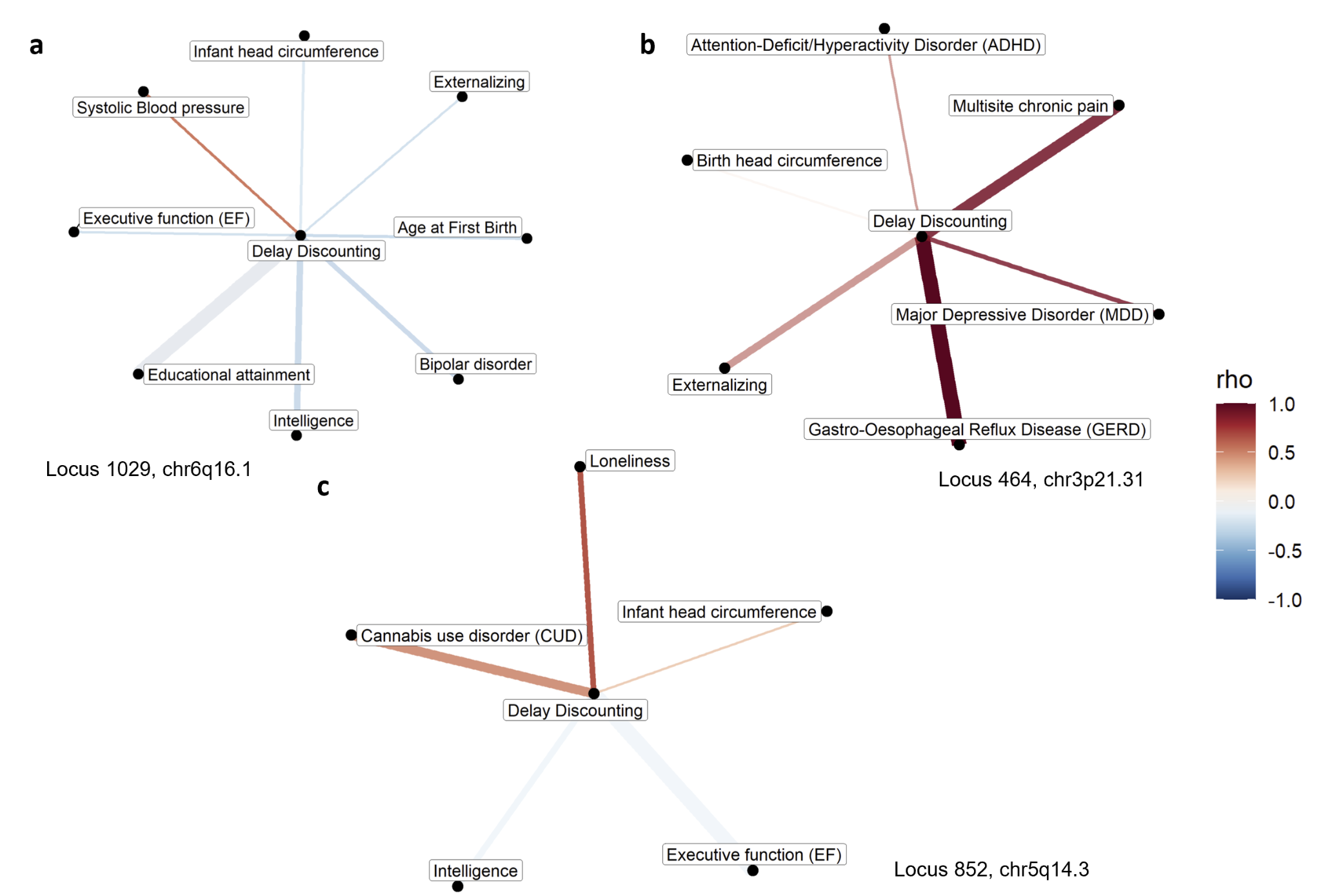

#### Supplementary Figure 16. Network plots showing the bivariate local genetic correlations within the hotspots a) locus1029, chr6q16.1 b) locus 464, chr3p21.31 and c) locus 852, chr5q14.3. Color of the edge indicates the direction of the local genetic correlation and line thickness indicates the significance.

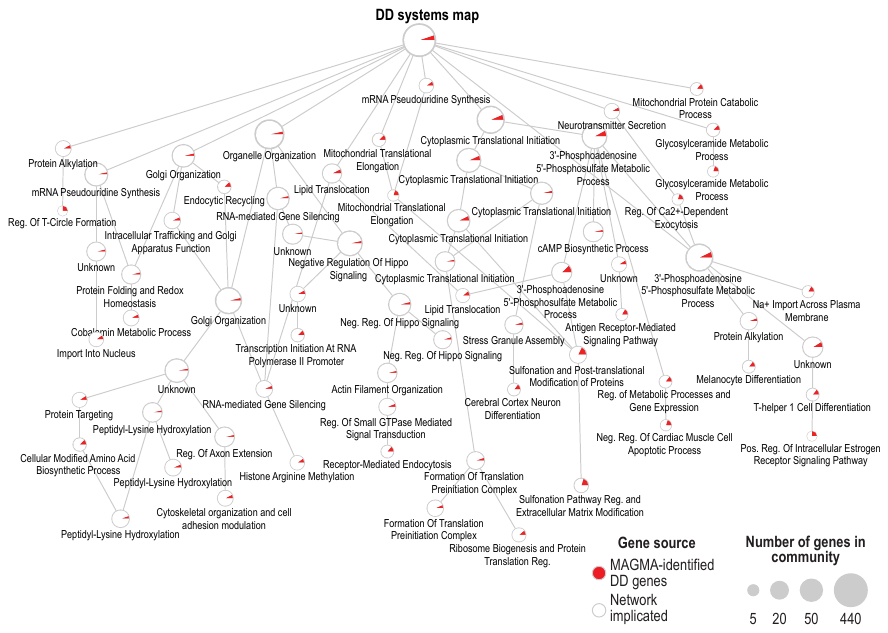

#### Supplementary Figure 17. System hierarchy for DD propagated from MAGMA-associated genes. For more detailed information on the biological processes, see Supplementary Table 14.

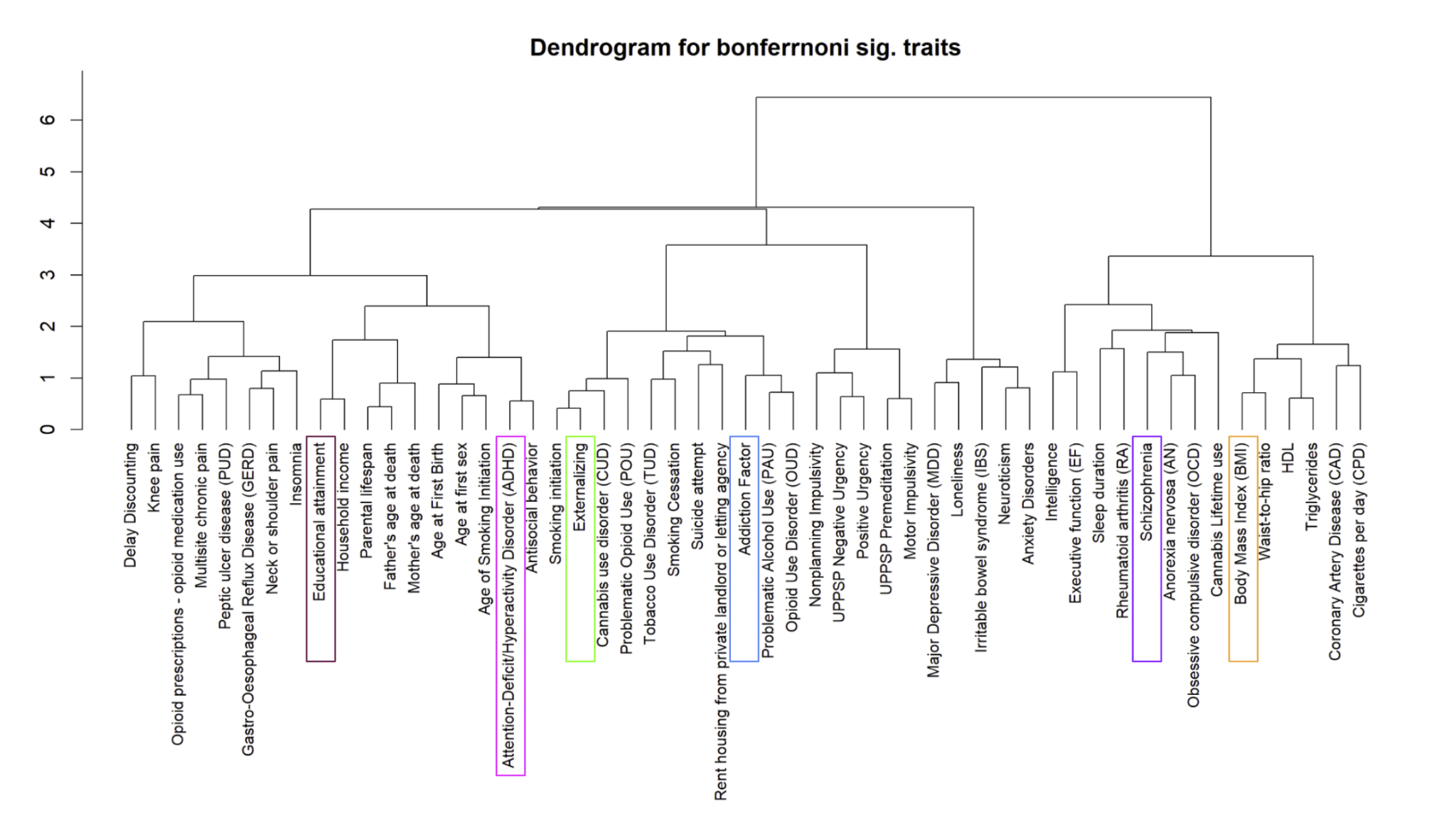

#### Supplementary Figure 18. Trait selection for network analysis based on dendrogram clustering of traits that were globally genetically correlated with DD at Bonferroni significance (Supplementary Table 12). Representative traits for clusters of interest that were used in the network analysis are highlighted.

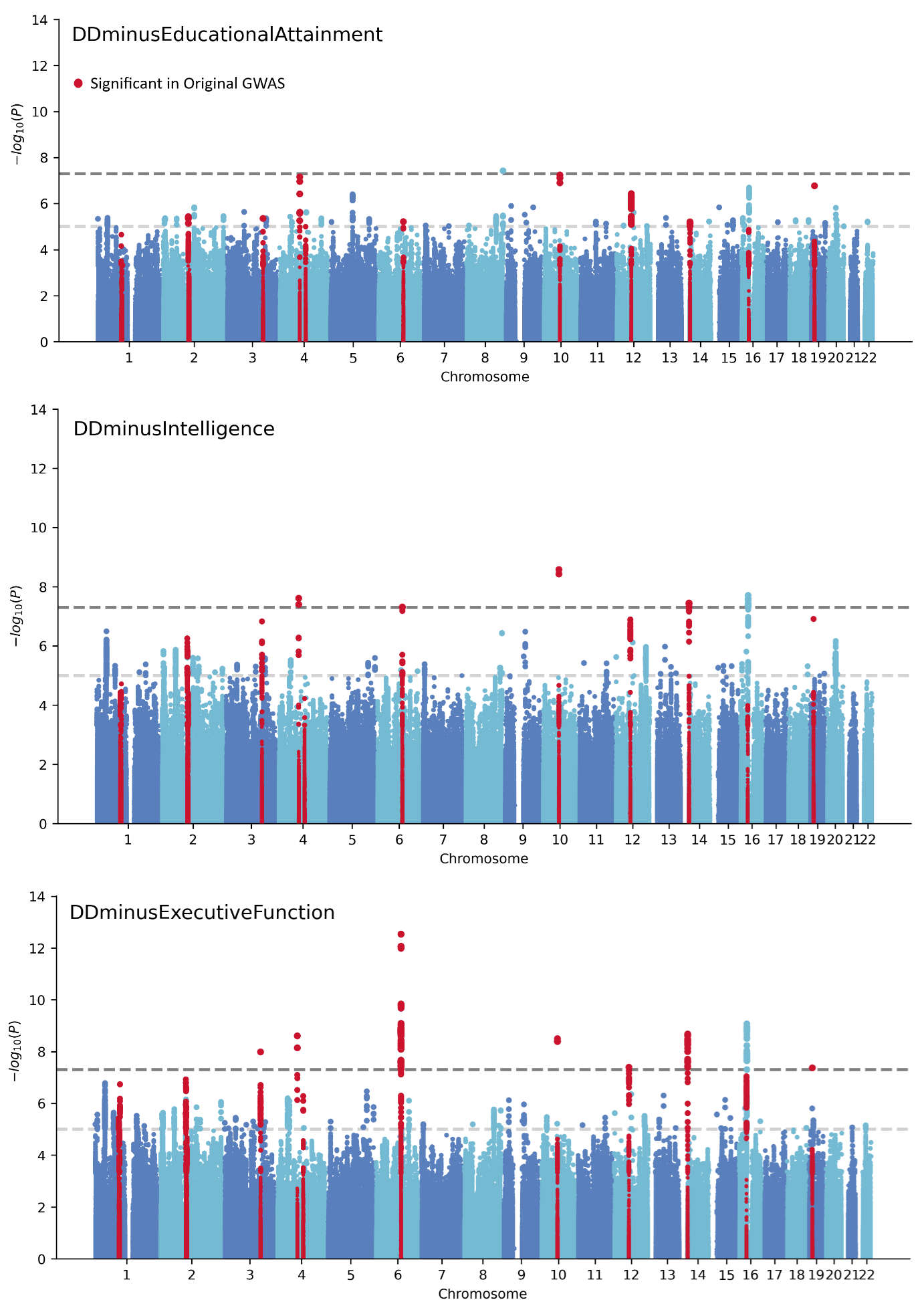

#### Supplementary Figure 19. Manhattan plot for the GWAS of DD after subtracting a) educational attainment^21^, b) intelligence^22^, and c) executive function^23^. The upper dashed line indicates genome-wide significance (*P* < 5.00x10^-8^), the lower dashed line indicates the threshold for suggestive associations (*P* < 1.00x10^-5^), and the red dots show SNPs that are significant (*P* < 5.00x10^-8^) in the original GWAS of DD.

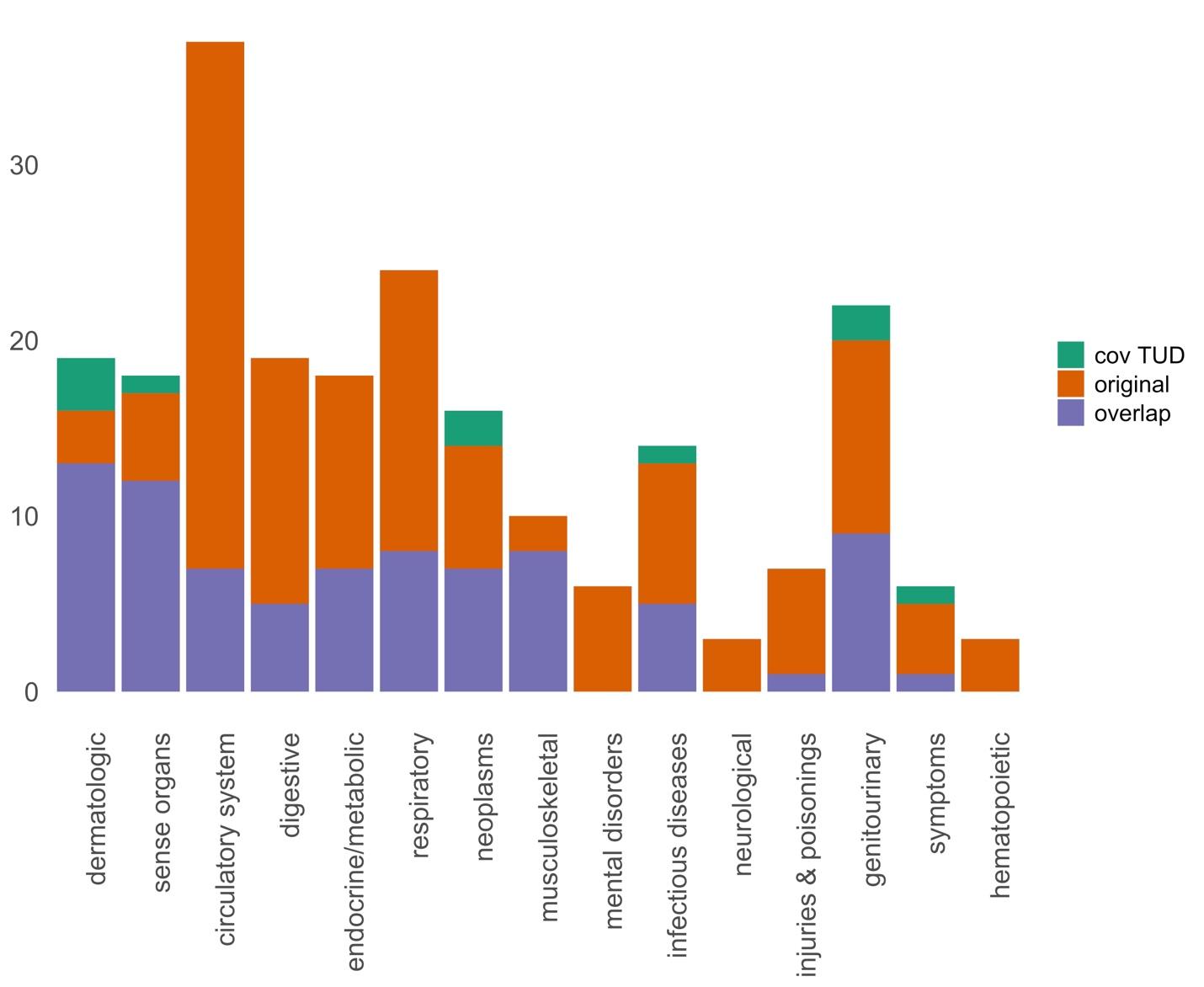

#### Supplementary Figure 20. Number of PheWAS associations by health category that were significant in both the original analysis and the analysis adjusted using tobacco use disorder as a covariate (cov TUD; purple), were significant only in the original analysis (orange), or were significant only in the cov TUD analysis (green).
